# Using routine primary care data to assess Sudden Cardiac Arrest risk in people with type 2 diabetes: a proof-of-concept case-control study

**DOI:** 10.1101/2024.01.30.24301990

**Authors:** Peter P. Harms, Laura H van Dongen, Nicky I.C. Oosterbaan, Frank C. Bennis, Joline WJ Beulens, Karin M.A. Swart, Mark Hoogendoorn, Hanno L. Tan, Petra P.J.M. Elders, Marieke T. Blom, RESCUED investigators

## Abstract

**Background:** Approximately 50% of out-of-hospital Sudden Cardiac Arrest (SCA) occurs in people with unrecognized SCA-risk and no preceding cardiologic care records. General practitioner (GP) records include these people, specifically people with type 2 diabetes (T2D) with increased SCA-risk. We aimed to provide a proof-of-concept for using routine primary care data to study SCA-risk in people with T2D.

**Methods:** This case-control study, identified SCA cases through the AmsteRdam REsuscitation STudies (ARREST) registry of out-of-hospital SCA in the Dutch region of Noord-Holland (2005-2019). We included cases with presumed cardiac cause and T2D registered at participating GP practices from the PHARMO Data Network and the academic network of general practice Amsterdam UMC (ANHA). Cases were matched (age, sex, T2D, GP-practice) with up to five non-SCA controls. From their GP files, we collected clinical measurements, medication use and medical history. Associations with SCA were analysed using univariable and multivariable conditional logistic regression (Odds Ratios, 95% confidence intervals).

**Results:** We included 247 cases and 1,143 controls. In the multivariable model, high fasting glucose (1.08 (1.01-1.16) per 1 mmol/L), high cholesterol ratio (1.17 (1.03-1.34)), moderate albuminuria (2.77 (1.84-4.16)), severe albuminuria (2.96 (1.44-6.08)), dyslipidaemia (0.53 (0.33-0.86)) and a history of cardiovascular disease (1.72 (1.23-2.17)) were significantly associated with SCA. Current smoking behaviour, decreased eGFR, insulin use, hypertension and microvascular complications were close to significantly associated with SCA.

**Conclusions:** The relatively strong associations in our small sample are consistent with those found in cardiologic care populations, indicating that GP file data can be useful to study SCA-risk.

## Introduction

Sudden cardiac arrest (SCA) is a substantial public health problem and the immediate cause of up to 50% of cardiac deaths and approximately 20% of all mortality in high-income countries^1,2^. In the European Union, out-of-hospital SCA incidence rates of 48-58 per 100.000 person years equate to an estimated 343.000 cases per year^3^. Only 6-20% of SCA victims survive^4^, and those that do can experience disabilities and enduring decline in general health^5^. Early recognition of SCA susceptibility could be advantageous for prevention of SCA.

Recognition of people that are vulnerable to SCA requires insight into predictors of increased SCA-risk. To date, research into predictors of SCA has often concentrated on people at increased SCA-risk that were often in treatment for cardiovascular disease (CVD), frequently coronary heart disease (CHD), by a cardiologist before the SCA, and their cardiologic care records. Beside rare arrhythmogenic genetic syndromes^6^, these efforts have already yielded a vast amount of literature on markers of SCA-risk such as prevalent diabetes, current smoking behaviour, dyslipidemia, hypertension, chronic kidney disease and a history of microvascular complications^7-14^. These are now established risk factors for out-of-hospital SCA and cardiovascular complications in general.

However, many studies indicate SCA-risk recognition remains challenging in the setting of CVD and CHD, especially in people with T2D^14-16^. Moreover, an out-of-hospital SCA is often the first manifestation of (coronary) heart disease^17,18^. Accordingly, around 50% of SCA victims have never consulted a cardiologist prior to the SCA, and are therefore not represented in cardiologic care records^19^. Furthermore, even when some of these SCA victims without prior CVD history are included in a study, for example in studies using death certificates or population cohorts, there is often insufficient data on cardiovascular risk factors or there are not enough SCA cases and thus not enough statistical power to study SCA^20^.

This dilemma might be resolved by studying medical files from general practitioners (GPs), since these also include people with unrecognized SCA-risk. Studying GP files with routine care data might provide useful novel clues for recognition of SCA-risk. However, the potential and validity of routine primary care data for studying SCA-risk must first be evaluated. A suitable population to do so would be people with type 2 diabetes (T2D) in primary care. Firstly, people with T2D have an over two-fold increased risk of SCA^11,15,20^. Moreover, in the Netherlands GPs invite people with T2D to regular check-ups to manage the T2D including the elevated risk of cardiovascular complications resulting in comprehensive GP records^21^.

Therefore, by assessing whether established SCA-risk factors measured in primary care are associated with SCA in people with T2D, we aimed to provide a proof-of-concept for using routine primary care data to study SCA-risk.

## Methods

### Sudden Cardiac Arrest cases

In this case-control study, SCA cases were identified through the AmsteRdam REsuscitation STtudies (ARREST) registry^22^. ARREST is a large scale, ongoing, and community-based registry which has prospectively included all out-of-hospital cardiac arrest resuscitation attempts by emergency medical services in the Dutch region of Noord-Holland since 2005. From 2007 onwards, an electrocardiogram (ECG) was used to document ventricular tachycardia or ventricular fibrillation during the resuscitation to confirm a cardiac cause of the cardiac arrest (as opposed to non-cardiac causes such as a ruptured aneurysm or a pulmonary embolism).

For the present study, we selected all out-of-hospital cardiac arrest resuscitation attempts with a presumed cardiac cause registered in ARREST between 2005-2019 involving residents over 18 years of age from the two respective catchment areas of participating GPs (Figure 1). Cardiac cause was presumed if emergency medical services had initiated a defibrillation procedure immediately after arrival at the emergency site. We excluded all resuscitation attempts that started in the ambulance during transport for non-emergency reasons or if the initial call for attendance of emergency medical services did not concern a potential resuscitation.

**Figure 1.**
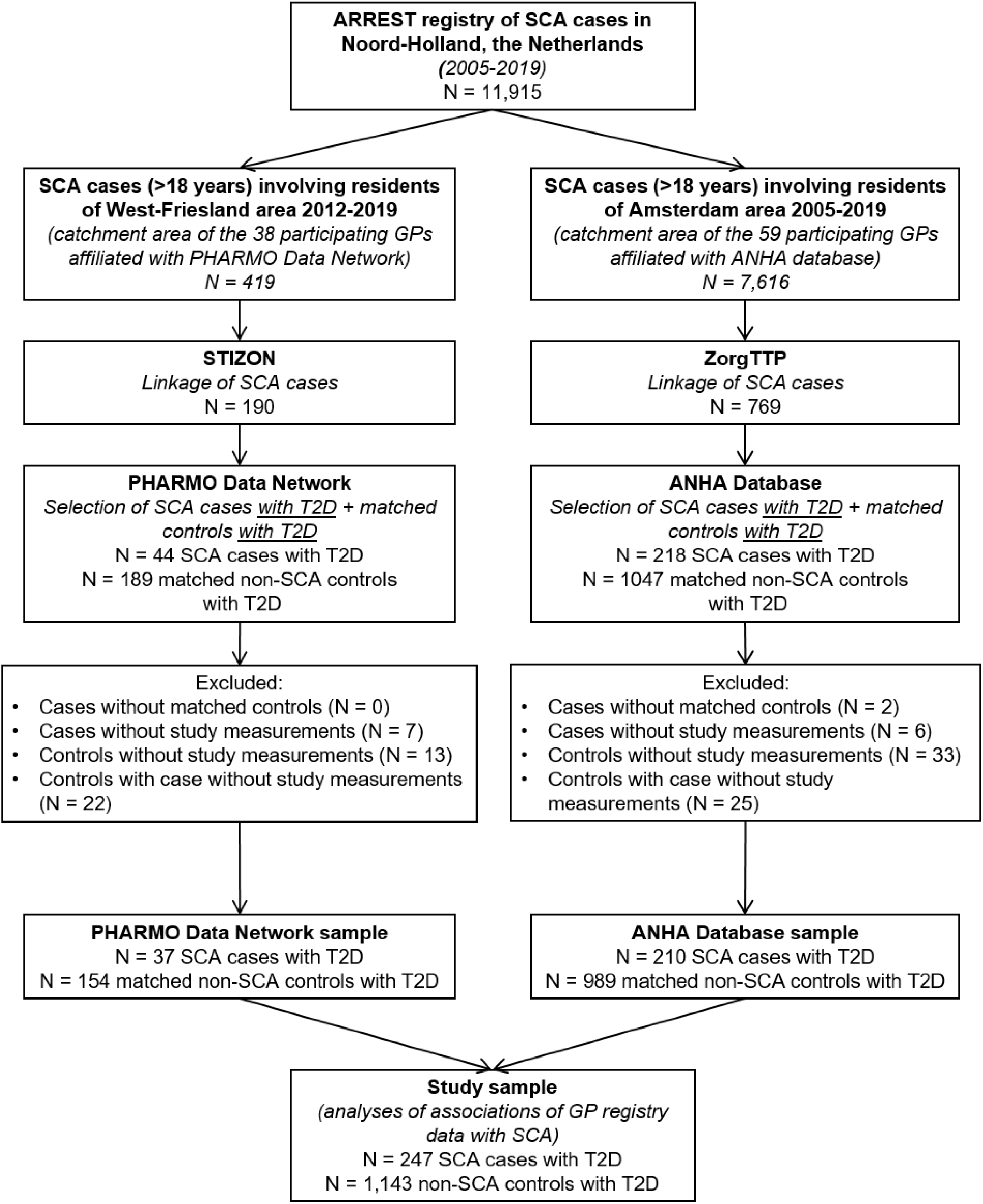
Flowchart of SCA cases, linkage to GP files and matching of controls.

### Linkage to GP files

We proceeded to link the selected SCA cases from ARREST to their GP electronic medical records in either the PHARMO Data Network^23^ or the academic network of general practice at Amsterdam UMC, location VUmc (ANHA database). The PHARMO Data Network includes GP practices from all over the Netherlands, of which (in 2020) 38 practices located in the West-Friesland area of the Noord-Holland region of the Netherlands agreed to participate in this study. ANHA is an ongoing longitudinal database containing routine general practice care data extracted from the electronic medical records of (in 2020) 59 participating practices in or near the city of Amsterdam. General practices in the PHARMO Data Network and ANHA provide their patients with information about the use of pseudonymised routine primary care data for research and quality of care purposes. Data from patients who object (opt-out) is excluded from use for research purposes.

To safeguard current ethical and privacy standards concerning medical research two Trusted Third Parties (TTP) executed the procedure linking the selected SCA cases from ARREST to their GP files. The Foundation for Information Provision for Care and Research (STIZON)^24^ performed the linkage with the PHARMO Data Network. The ZorgTTP foundation (https://www.zorgttp.nl/) performed the linkage with the ANHA database.

### Type 2 Diabetes

We assessed if SCA cases had T2D prior to the SCA by considering both the medical history prior to the event registered in ARREST, and captured in their electronic GP patient file. T2D was defined as either a history of T2D reported to ARREST by the GP of the SCA case, or a history of T2D according to the GP patient file. GP file history of T2D was operationalised as the presence of an International Classification of Primary Care (ICPC) code for T2D (T90.02), or alternatively an ICPC code for diabetes (T90.00 or T90) without presence of type 1 diabetes code (T90.01), or any prescription of glucose lowering medication that is not insulin (Anatomical Therapeutic Chemical code A10B).

In general, Dutch GPs register a T90.02 code based on a T2D diagnosis according to the criteria by the Dutch College of General Practitioners (NHG)^25^. These criteria are either two fasting glucose measurements ≥ 7.0 mmol/L on two different days or one fasting glucose measurement 7.0 ≥ mmol/L or plasma glucose measurement ≥ 11.1 mmol/L in combination with symptoms that match hyperglycemia.

### Matched Controls

After linking SCA cases to their GP data, cases with T2D were matched with up to five (when available) non-SCA controls with T2D from the same GP practice. More precisely, controls had to be diagnosed with T2D according to the above mentioned criteria regarding electronic GP patient file information, and registered at the same GP practice, during the time period before the SCA date of the case. Controls were additionally matched based on sex (male/female), and age (maximum difference of five years). To prevent a control from being matched with multiple cases, selection and case allocation was guided by match rank numbers based on smallest age difference, and the restriction that a potential control was matched to one case only.

### Collection of routine primary care data

After matching, the relevant data of cases and controls was extracted from the electronic medical records stored in the PHARMO Data Network and ANHA database. The data was extracted from the clinical measurement file (Dutch College of General Practitioners (NHG) codes), the medication prescriptions file (ATC codes), and disease episodes file (ICPC codes). Extraction was restricted to a maximum period of 5 years preceding the SCA date of the case, except for disease episodes to obtain complete medical history. For a summary of the selected NHG, ATC or ICPC codes please see Supplementary Table S1.

### Clinical measurements

The following clinical measurement data was extracted from the electronic GP files: Smoking behaviour (never, former smoker, and current smoker), Body mass index (BMI, in kg/m^2^), Systolic and diastolic blood pressure (mmHg), pulse frequency (beats/minute), HbA1c (mmol/mol), fasting blood glucose (mmol/L), total cholesterol (mmol/L), HDL-cholesterol (mmol/L), LDL-cholesterol (mmol/L), total cholesterol/HDL-cholesterol ratio, triglycerides (mmol/L), blood creatinine (umol/L), urine creatinine (mmol/L), urine albumin (mg/L), estimated glomerular filtration rate (eGFR) calculated by the Chronic Kidney Disease Epidemiology Collaboration (CKD-EPI) equation or alternatively the Modification of Diet in Renal Disease (MDRD) equation^26^ and urinary albumin creatinine ratio (UACR). eGFR was categorised as normal or high (>90 ml/min), mildly decreased (60-90 ml/min), moderately decreased (30-60 ml/min) or severely decreased (<30 ml/min). Albuminuria was categorised as normal to mild (<3 mg/mmol), moderate (3-30 mg/mmol) or severe (>30 mg/mmol). We analysed the clinical measurements that most closely preceded the SCA to evaluate the short term associations.

### Medication use and medical history

Medication use was categorised as present if at least one prescription was registered in the extracted electronic GP file data. Glucose-lowering medication (ATC: A10 codes) was classified as no medication, oral medication only (A10B), insulin use only (A10A), and combined oral and insulin use (A10A and A10B). Further medication use was categorised as antihypertensive medication (C02, C03 and C07-9 codes), lipid-lowering medication (C10 codes) or heart rate-corrected QT interval (QTc) prolonging medication which included antiarrhythmics (C01B codes), antipsychotics (N05AX codes) and antidepressants (N06A codes).

Medical history of hypertension was defined as a clinical measurement of elevated blood pressure (systolic>140 mmHg or diastolic>90 mmHg), antihypertensive medication use and/or a registered disease episode of hypertension (ICPC: K85-87 codes). Dyslipidemia included elevated LDL-cholesterol (>2.6 mmol/l), lipid-lowering medication use and/or a disease episode of dyslipidemia (T93 codes). Pre-existing cardiovascular disease was defined as a disease episode of coronary heart disease (K74, K75 and K76 codes), heart failure (K77 codes), transient ischemic attack (K89), stroke (K90 codes), atherosclerosis (K91) or intermittent claudication (K92.01). A history of microvascular complications comprised neuropathy (N94 and N94.02 codes), retinopathy (F83 codes) or chronic ulcers (S97 codes).

### Data cleaning, missing values and multiple imputation

Of the total 262 linked SCA cases with T2D, 2 (0.8%) were excluded because no matched non-SCA controls were available, and 13 (5.0%) were excluded because no information on the abovementioned clinical characteristics was present in their electronic GP files. The remaining 247 (94.3%) SCA cases with T2D were included in the analyses. Of the total 1236 matched non-SCA controls with T2D, 46 (3.7%) were excluded because there was no information on the abovementioned clinical characteristics in their electronic GP files, and 47 (3.8%) were excluded because they were matched with one of the 13 SCA cases that were excluded because there was no information on the clinical characteristics the electronic GP files. The remaining 1,143 (92.5%) controls with T2D were included in the analyses (Figure 1; summation of numbers from both the PHARMO Data Network and ANHA database).

Measurements of continuous variables with values outside the physically possible range were replaced by the previous closest measurement in time or considered missing if no alternative measurement was available. Variables with (historically) different units of measurement were converted to the current standard unit, for example HbA1c% was converted to mmol/mol using the formula recommended for Dutch GPs^27^.

After data collection and cleaning, 498 (35.8%) of the SCA cases and matched non-SCA controls had complete data on all variables: age, sex, the 4 medication use, the 4 disease history and the 17 clinical measurement variables. Missing values were present only in the clinical measurement variables (Table 1). The number of missing clinical measurement values per participant ranged from 0 to 17 and the distribution was skewed to the left with a median (IQR) of 2 (9). Notably, 15.3% of participants had missing values for all 17 clinical measurement variables. These missing values were imputed using only the participant’s age, sex, medication use, and disease history.

**Table 1.** Distribution of number of missing values for clinical measurements per participant.

| Number of missing clinical measurements values per participant | Number of participants | Percentage (%) |
| --- | --- | --- |
| 0 | 498 | 35.8 |
| 1 | 187 | 13.5 |
| 2 | 81 | 5.8 |
| 3 | 74 | 5.3 |
| 4 | 56 | 4.0 |
| 5 | 65 | 4.7 |
| 6 | 30 | 2.2 |
| 7 | 16 | 1.2 |
| 8 | 29 | 2.1 |
| 9 | 20 | 1.4 |
| 10 | 19 | 1.4 |
| 11 | 18 | 1.3 |
| 12 | 16 | 1.2 |
| 13 | 19 | 1.4 |
| 14 | 17 | 1.2 |
| 15 | 27 | 1.9 |
| 16 | 6 | 0.4 |
| 17 | 212 | 15.3 |
| Total | 1390 | 100.0 |

Proportions of missing values per clinical measurement variable (range: 19-50%) were consistently somewhat higher in SCA cases compared with non-SCA controls (Table 2). Further missing value analyses showed a variety of different missing value patterns (n=175), and that some of the missingness was connected with non-missing values in other variables, indicating that values were partially “missing at random” (MAR) and not all “missing not at random” (MNAR) or “missing completely at random” (MCAR)^28-31^ (Supplementary Table S3.1-S3.12).

**Table 2.**
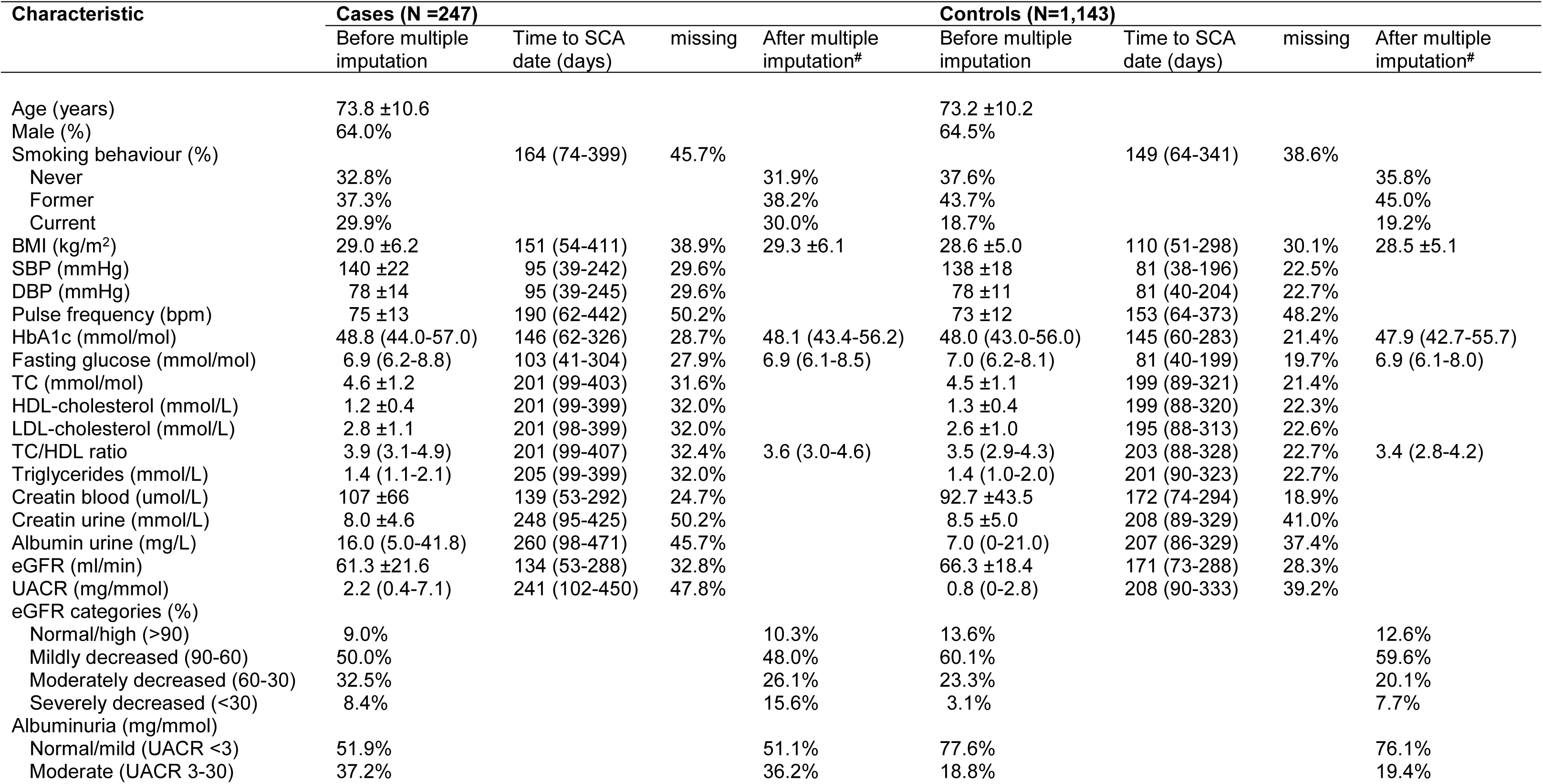
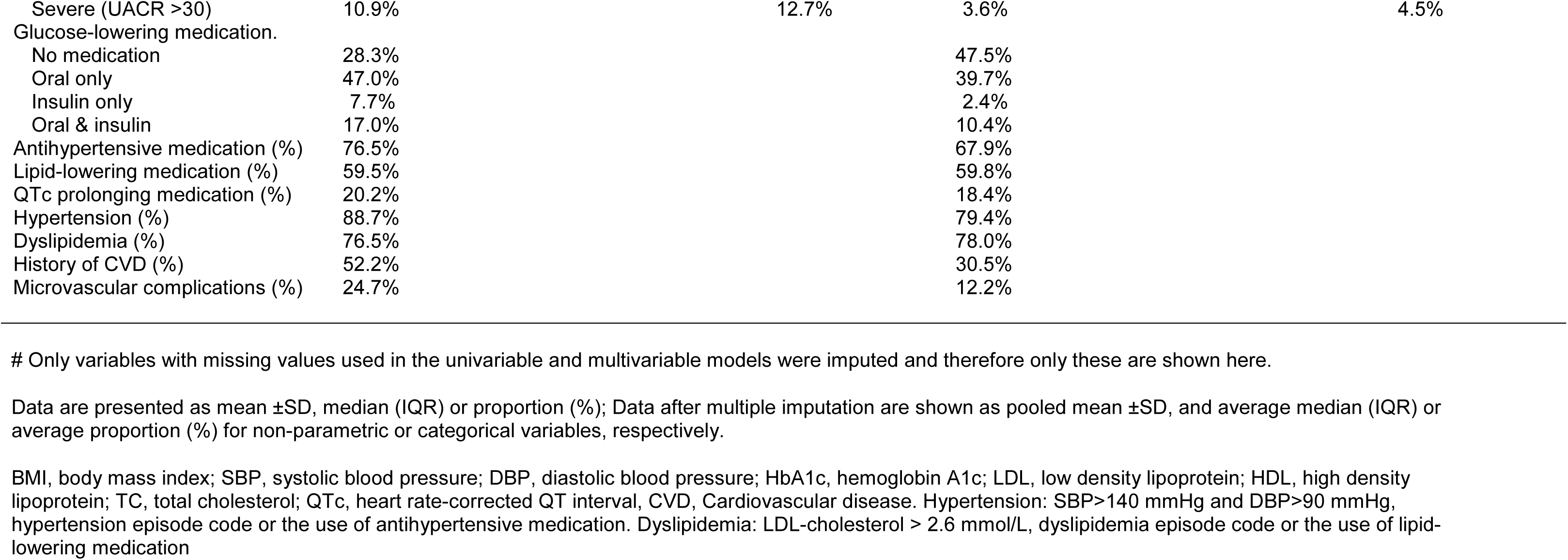
Study population characteristics before and after multiple imputation, stratified for SCA cases with T2D and matched non-SCA controls with T2D.

Missing values were imputed using Multiple Imputation by Chained Equations (MICE), with predictive mean matching for continues variables, logistic regression for dichotomous variables, polytomous regression for categorical variables, 50 iterations and 50 imputed datasets^28-31^. We checked absence of multi-collinearity (correlation coefficients <0.80) and successful convergence (spaghetti-plots) of the imputation model. The imputation model included all variables of the analyses model (see below), the outcome variable (case/control) and informative auxiliary variables (age, sex, SBP, DBP, total cholesterol, HDL-cholesterol, triglycerides, blood creatinine, anti-hypertensive and lipid-lowering medication). Albuminuria and smoking behaviour were not used as predictors in de imputation model because proportion of missing was >40%.

### Statistical analyses

Successful matching was checked by pairwise comparison of age, sex, T2D status and GP practice number for each case and its matched controls (Supplementary Table S2). Study population characteristics were calculated for cases and controls in means (±SD), medians (IQR) or proportions (%) as appropriate.

Univariable and multivariable conditional logistic regression models were used to assess the association of clinical characteristics with SCA, computing odds ratios (ORs) with 95% confidence intervals (95%CIs) for smoking behaviour, BMI, HbA1c, fasting glucose, cholesterol ratio, eGFR category, Albuminuria, glucose-lowering medication, QTc-prolonging medication, hypertension, dyslipidemia, history of CVD, and microvascular complications. The multivariable model included all variables simultaneously. A sensitivity analyses compared conditional and unconditional (adjusted for matching variables sex and age) logistic regression models because it is debated which is most appropriate for matched case-control studies^32^. These analyses were conducted with the imputed datasets. In a second sensitivity analysis, the influence of multiple imputation was evaluated by comparing the univariable associations with SCA from the original dataset (complete cases and controls) and the imputed datasets (pooled estimates) for all variables with missing values. Assumptions of binary logistic regression analysis were checked for all analyses.

Pooled estimates or p-values of the imputed datasets are presented when possible, otherwise the average estimate or p-value over the imputed datasets is given^33^. For all analysis, p-values of <0.05 were regarded as significant (two-sided). Analyses were performed with SPSS version 26 (IBM corporation, New York, USA) and R (studio) version 4.0.3 (R foundation for statistical computing, Vienna, Austria, http://www.r-project.com) in combination with the R packages *tidyverse* (1.3.2), *table1* (1.2.4), *expss* (0.11.1), *mice* (3.14.0), *miceadds* (3.13.12), and *survival* (3.3.1).

## Results

### Study population characteristics

The 247 SCA cases with T2D had a mean age of 73.8 ±10.6 years, and 64.0% was male. The 1,143 matched non-SCA controls with T2D had a mean age of 73.2 ±10.2, and 64.5% were male. The median (IQR) time between the clinical measurements and the SCA date of the case was also comparable for cases and controls, and ranged from 95 (39-242) days for SBP to 260 (98-471) days for urinary albumin in cases, and from 81 (38-196) days for SBP to 208 (90-333) days for UACR in controls. After multiple imputation, the descriptive statistics of the imputed variables were comparable with those of the original data, indicating imputation was successful (Table 2).

### Associations with sudden cardiac arrest

Current smoking, higher fasting glucose, higher cholesterol ratio, severely decreased eGFR, moderate and severe albuminuria, insulin only use, no dyslipidemia, hypertension, and a history of CVD or microvascular complications were significantly associated with SCA in the univariable analyses (Figure 2). After adjustment for the other clinical characteristics in the multivariable analysis, high fasting glucose (OR 1.08 95%CI (1.01-1.16) per 1 mmol/L), high total cholesterol/HDL-cholesterol ratio (1.17 (1.03-1.34)), moderate albuminuria (2.77 (1.84-4.16), severe albuminuria (2.19 (1.44-6.08)), and a history of cardiovascular disease (1.68 (1.26-2.24) or microvascular complications (1.48 (1.02-2.16)) remained significantly associated with SCA. Dyslipidemia (0.53 (0.33-0.86)) also remained inversely associated in the multivariable analysis. Current smoking behaviour (1.62 (0.99-2.71)), severely decreased eGFR (2.19 (0.94-5.08)), insulin use (1.97 (0.88-4.39)), hypertension (1.33 (0.79-2.25)) and microvascular complications (1.44 (0.96-2.17)) were almost significantly associated with SCA in the multivariable analyses. BMI, HbA1c and QTc-prolonging or oral glucose lowering medication use were not associated with SCA in both the univariable and multivariable analyses.

**Figure 2.**
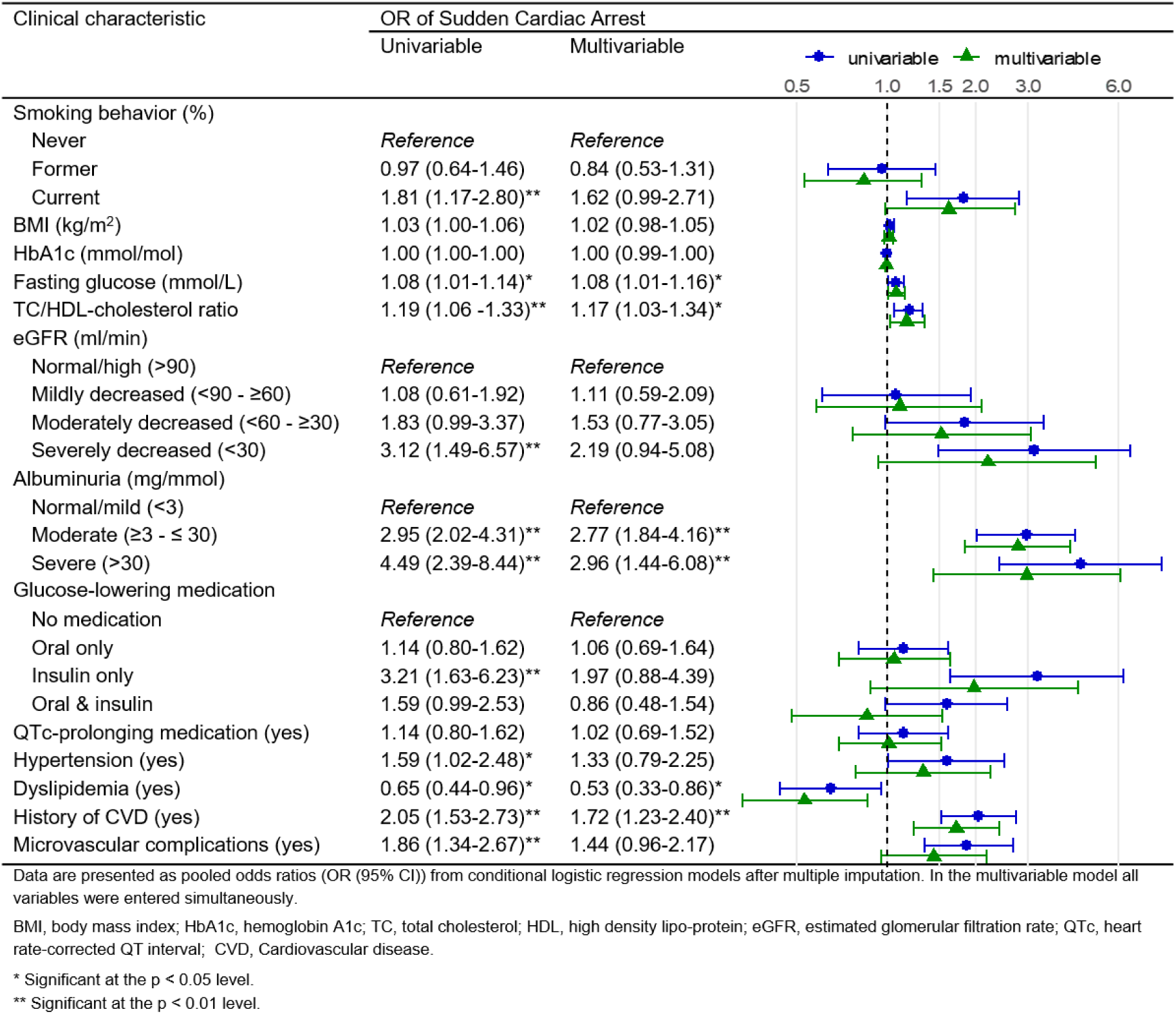
The association of clinical characteristics with SCA in people with T2D, comparing SCA cases with matched non-SCA controls.

In the sensitivity analyses, the associations with SCA were similar to those from main analyses (Supplementary Table S4 and Supplementary Table S5).

## Discussion

This proof-of-concept study assessed the usefulness of routine primary care data for the study of SCA-risk by evaluating whether established SCA-risk factors measured in primary care are indeed associated with SCA in people with T2D. Known SCA-risk factors such as a clinical history of macro- or microvascular disease (i.e. CVD, microvascular complications, decreased eGFR, albuminuria) or its major risk factors (i.e. smoking, hypercholesterolemia and hypertension) were associated with an up to two-fold increased risk of SCA in persons with T2D. Additionally, a 1 mmol/L higher fasting glucose increased SCA-risk with nearly 10%.

### Main findings in perspective

To our knowledge, no previous studies investigated associations of clinical characteristics with SCA in people with T2D using routine primary care data. However, a case-control study in King County, Seattle (USA) on pharmacologically-treated people with T2D in ambulatory care tested only for univariable differences between some of the same clinical characteristics^14^. Like this current study, that study also reported significant univariable associations of smoking behaviour, insulin treatment, and a history of cardiovascular disease or microvascular complications with SCA.

A history of CVD, specifically CHD,^1,2^ or microvascular complications^11-14^ are well established SCA-risk factors that usually convey an over 50% increased relative risk. Consistently, we report approximately the same one-and-a-half-fold increased SCA-risk for people with T2D and a history of CVD or microvascular complications in primary care. Moreover, there is a vast amount of literature on SCA-risk in people with T2D and CVD, specifically CHD, in which smoking behaviour, hypertension, dyslipidemia, and nephropathy are consistently associated with increased SCA-risk^7-16^. It would be beyond the scope of this paper to review all this previous literature on these well-known associations. However, below we highlight some reports on these SCA-risk markers in people with T2D and CVD, specifically CHD, that are relevant to this study’s aim. This study aimed to provide a proof-of-concept that primary care data are a valid data source which includes both SCA victims with and without a CVD or CHD history prior to the SCA event, by showing that known risk factors for SCA in people with CVD or CHD are indeed also present and associated with SCA in primary care data.

Chronic kidney disease has long been recognised as a strong SCA-risk factor^9,10^. Therefore, the approximately twofold increased risk of SCA for people with severe or moderate albuminuria in our proof-of-concept study indicates validity of using routine primary care data in SCA-risk research. Additionally, it is also consistent with a recent report from the prospective population-based REGARDS study^34^ that found associations (hazard ratio, 95%CI) for moderate (1.53 (1.11-2.11)) and severe (1.56 (1.17-2.11)) albuminuria over a median follow-up of 6.1 years. Moreover, the REGARDS study concluded albuminuria is a more important predictor of SCA-risk than eGFR because only severely decreased eGFR (<45 mL/min/1.73 m(2)) was significantly associated with SCA (1.66, 1.06-2.58). Even though the associations with SCA we report for eGFR are probably non-significant due to our small sample size, their similar magnitude is compatible with the findings from the REGARDS study.

Smoking is a modifiable risk factor for all kinds of CVD, including SCA. Accordingly we also found a firm association of current smoking with SCA. Additionally, the absence of an association of former smoking with SCA underlines the findings of a systematic review and meta-analysis in people with T2D that showed smoking cessation reduces the risk of CVD^35^. A similar rationale extends to the association of high total/HDL cholesterol ratio and simultaneous inverse association of dyslipidemia with SCA: treatment for dyslipidemia with statins effectively lowers the cholesterol ratio by lowering total/LDL cholesterol, with subsequent lower risk cardiovascular complications such as SCA. Hypertension is another well-established SCA-risk factor that also in this study was associated with an over 30% increased relative SCA-risk^7,8^. Additionally, one other SCA study that used routine primary care data was conducted in people with hypertension, and it showed that despite prescribed antihypertensive medication there still remained an increased SCA-risk comparable to this study^36^.

While an increased fasting glucose was associated with a slightly increased odds of SCA, HbA1c was not associated with SCA in our study. As the International Hypoglycemia Study Group discussed in a recent review, hypoglycemia events are associated with increased risk for CVD events and mortality, but this is not reflected in an association of low HbA1c with CVD events and mortality^37^. Increased fasting glucose might be associated with SCA through its detrimental microvascular effects or as an indicator of metabolic changes.

### Future research

This proof-of-concept study validates the suitability of routine primary care data from people with T2D for the study of SCA, and the rationale of the RESCUED project^38^. Future studies with routine primary care data from people with T2D might provide useful novel clues for recognition of SCA-risk (in people with T2D) since GP contacts increase in the weeks before SCA^39^. In addition to the apparent cardiovascular and SCA-risk factors we analysed, GP files contain a broad scope of data on physical examinations, laboratory results, prescribed medication, medical history, free text consultation notes and meta-data like health services usage, communications and declarations. This longitudinal and prospectively recorded data from before SCA events offer a profusion of possibilities to discover predictors of SCA that have until now remained unrecognised. Longitudinal analyses methods like GEE and mixed/multi-level modelling, latent class analyses or joint-modelling could elucidate risk factor, medication and disease trajectories that predispose to SCA liability. The application of machine learning techniques from non-medical data science domains could reveal complex and multifactorial patterns of clinical characteristics and health services usage that predict SCA vulnerability. Text mining analyses could harvest the nuances of context related professional experience and intuition that GPs incorporate into their free text consultation notes. Moreover, linkage and integration of routine (primary) health care data with social demographic, environmental, metabolic and genetic data would create opportunities for multidimensional approaches.

### Strengths and limitations

This study assessed whether routine GP care data can be useful when studying SCA-risk. Routine care data studies typically suffer from missing data issues. It is sometimes argued that multiple imputation should not be used to handle (large proportions) of missing data. However, multiple imputation mitigates the problems that arise with complete case analyes^30^, even for large proportions of missing data^31^. Complete case analyses lead to biased results when the missingness is not completely at random. This is often the situation in routine care because measurements are performed only when indicated for the management of disease. The similar results from the univariable sensitivity analyses (complete case vs multiple imputation) suggest this did not seriously affect our results. Nevertheless, complete case analyses or multiple imputation is not a panacea for missing data and the results should be interpreted with caution.

Another strength of this study is the SCA case definition from the ARREST cohort where out-of-hospital SCA was assessed by emergency medical services during the event, often with the aid of an ECG. In addition, the case-control design is vulnerable to selection bias due to different a-priori probability of study inclusion for cases and specifically controls^40^. We alleviate this by including people with T2D that regularly visit their GP for routine check-ups. That only about 7% of the linked cases and matched controls had to be excluded indicates the GP records from people with T2D were comprehensive and that there was little selection. Moreover, we used ∼5 matched controls from the same GP practice and calendar time period to control for confounding by age, sex, diabetes, care provider differences and time trends and used unconditional and conditional multivariable regression models.

Causation should not be readily inferred because of the observational study design and because the measurements of the determinants were taken a median six months before the SCA event. Lastly, healthcare registry measurements are not collected according to a strict study protocol that minimises measurement error. However, Dutch GPs perform physical examinations according to standards of the Dutch College of General Practitioners (NHG) and order laboratory test at accredited clinical laboratories.

## Conclusions

The relatively strong associations of established SCA-risk factors with SCA observed in our small sample of people with T2D from primary care are consistent with previous literature on SCA-risk factors in cardiologic care populations. This indicates that GP files of people with T2D can be useful to study SCA-risk and more extensive future studies might well aid the discovery of novel SCA-risk predictors.

## Acknowledgements

This study has been made possible by collaboration with the ARREST study, the Foundation for Information Provision for Care and Research (STIZON), the ZorgTTP foundation, the PHARMO Institute, and the academic network of general practice Amsterdam UMC (ANHA). The authors thank participants of the ARREST study as well as the collaboration partners. From ARREST: the authors thank C.M. de Haas, V. van Eeden, R. Stieglis and L.A.E. Bijman for data management, and R.W. Koster for management of the ARREST project. Moreover, the authors are greatly thankful for all the students for collecting data, the participation of all EMS dispatch centers (Amsterdam, Haarlem, Alkmaar), regional ambulance services (Ambulance Amsterdam, GGD Kennemerland, Witte Kruis, Ambulancezorg Veiligheidsregio Noord-Holland Noord), fire brigades, and police departments, as well as general practitioners and hospitals in the study region. From STIZON and ZorgTTP: the authors thank all involved staff for judiciously linking the ARREST participants with their GP electronic healthcare records. From the PHARMO Institute and the ANHA: the authors thank everybody involved in building, managing and maintaining these databases. Special thanks to Hanna H.K. Joosten and Pauline Slottje from the ANHA for their assistance with the (privacy legislation related) practicalities of data amalgamation, development of the algorithm used for the matching of cases and controls, and assembly of the raw ANHA datasets used for this study.

## Ethics declaration

The ARREST study was approved by the medical ethical review committee (METC) of the Amsterdam UMC, location AMC (decision no.: 2017_260), and conducted according to the Dutch Medical Research Involving Human Subjects Act (WMO), which is in-line with the principles of Good Clinical Practice (GCP) and the declaration of Helsinki. Written informed consent was obtained from all cases that survived SCA, after discharge from the hospital. The use of data from SCA victims that did not survive, and were therefore unable to give consent, is permitted by the Dutch WMO Act. The PHARMO Data Network and ANHA database contain pseudonimysed routine primary care data from all patients enlisted in the participating general practices, except for patients who object to the use of their data for research and quality of care purposes. The medical ethical review committee (METC) of the Amsterdam UMC, location VUmc (decision no.: 2018.579), confirmed that the current study including routine primary care data from non-SCA controls is not subject to the WMO act 2018.

## Data availability

In compliance with Dutch and European privacy legislation, the routine primary care registry data that support the findings of this study were used under strict license for the current study only, and so are not publicly available.

## Author contributions

PPH, LHvD, NICO, JWJB, MH, HLT and PE, MTB contributed to conception, design, data acquisition and analyses, and interpretation of the results. FCB and KMAS contributed to data acquisition. PPH drafted the first manuscript. All authors reviewed the first and subsequent versions. All authors agreed to be accountable for aspects pertaining integrity or accuracy, and approved the final version. PPH, MTB and PE are the guarantors of this work, and as such, had full access to all the data in the study and take responsibility for the integrity and the accuracy of the data analysis.

