## Supplementary Table S for "Using routine primary care data to assess Sudden Cardiac Arrest risk in people with type 2 diabetes: a proof-of-concept case-control study"

#### Supplementary Material

##### Contents

- Table S1. Coding system and codes used to define the clinical characteristics.
- Table S2. Distribution and differences in matching criteria for cases and controls.
- Table S3.1-S3.12. Association of missingness with non-missing values in other variables.
- Table S4. Sensitivity analysis comparing associations with SCA from conditional and unconditional (adjusted for sex and age) logistic regression models.
- Table S5. Sensitivity analyses assessing the influence of multiple imputation by comparing the univariable associations with SCA from original and imputed datasets.

Table S1: Coding systems and codes used to define the clinical characteristics.

| Characteristic | Coding system | Code |
|---|---|---|
| Clinical measurements | NHG | |
| Smoking behavior | | 1739 |
| Weight | | 357, 2408 |
| Height | | 560 |
| BMI | | 1272, 1234 |
| SBP | | 1744, 2055, 2659, 2668, 3326, 3336, 1794 |
| DBP | | 1740, 2056, 2660, 2669, 3327, 3337 |
| Pulse frequency | | 1875, 3320, 3694, 3963 |
| HbA1c | | 2816, 368, 2645, 415, 416 |
| Fasting glucose | | 372, 382, 3710, 3208 |
| TC | | 192, 3852 |
| HDL-cholesterol | | 446, 3853 |
| LDL-cholesterol | | 542, 2683, 3854 |
| **TC/HDL-cholesterol ratio** | | 181, 3857 |
| Triglycerides | | 1377, 3855 |
| eGFR | | 3583, 3741, 3907, 3908, 1919, 3740, 1918, 524 |
| Creatinine urine | | 525, 526, 527 |
| Creatinine blood | | 523, 3739 |
| Albumin urine | | 38, 39 |
| UACR | | 40, 41, 42 |
| Medical prescriptions | ATC | |
| Glucose lowering medication excl. insulin | | A10B |
| Insulin or analogue | | A10A |
| Lipid-lowering medication | | C10 |
| Antihypertensive medication | | C02, C03, C07, C08, C09 |
| Diuretics | | C03 |
| Beta blocking agents | | C07 |
| Calcium channel blockers | | C08 |
| Agents acting on the renin-angiotensin system (ACE-inhibitors & ARB blockers) | | C09 |
| QTc-prolonging medication | | C01B, N05A, N06A |
| Antiarrhythmic | | C01B |
| Antipsychotics | | N05A |
| Antidepressants | | N06A |
| Medical history | ICPC | |
| Type 2 Diabetes | | T90.02 |
| Hypertension | | K85, K86, K87 |
| Dyslipidemia | | T93 |
| History of CVD | | K74, K75, K76, K77, K89, K90, K91, K92 |
| Coronary heart disease | | K74, K75, K76 |
| Angina pectoris | | K74 |
| Acute myocardial infarction | | K75 |

| | |
|---|---|
| Other ischemic heart disease | K76 |
| Heart failure | K77 |
| Transient Ischemic Attack | K89 |
| Stroke | K90 |
| Atherosclerosis | K91 |
| Intermittent claudication | K92 |
| Microvascular complications | |
| Neuropathy | N94 |
| Retinopathy | F83 |
| Chronic ulcers | S97 |

NHG: Nederlands Huisarts Genootschap (Dutch College of General Practitioners); ICPC: International Classification of Primary Care; ATC: Anatomical Therapeutic Chemical. BMI, body mass index; SBP, systolic blood pressure; DBP, diastolic blood pressure; HbA1c, hemoglobin A1c; LDL, low density lipoprotein; HDL, high density lipoprotein; TC, total cholesterol; QTc, heart rate-corrected QT interval, CVD, Cardiovascular disease.

Websites of the coding systems with background information:

- NHG diagnostic determinations: https://bepalingen.nhg.org/labcodes/determinations
- ICPC: https://www.nhg.org/themas/artikelen/icpc-online and https://www.globalfamilydoctor.com/groups/workingparties/wicc.aspx
- ATC: https://www.whocc.no/atc_ddd_index/

Table S2. Distribution and differences in matching criteria for cases and controls.

| Criterion | Cases (n=247) | Controls (n=1143) | Difference for case-control pairs |
|---|---|---|---|
| T2D (%) | 100 | 100 | 0 |
| GP practices (n) | 78 | 78 | 0 |
| Male (%) | 64 | 64 | 0 |
| Age at OHCA (mean ±SD) | 74 ±11 | 73 ±10 | Median 1, IQR 0-2, min 0, max 5 |

#### Table S3.1 - Separate variance t tests[a] by missingness indicators

| | | Age | BMI | SBP | DBP | HR | HbA1c | FG | TC | HDL | LDL | TC/HDL | TG | Creat B | Creat U | Alb U | eGFR | UACR |
|---|---|---|---|---|---|---|---|---|---|---|---|---|---|---|---|---|---|---|
| BMI | t | 1.6 | . | -2.2 | -2.4 | -.9 | -1.2 | -.6 | -4.7 | -1.8 | -3.6 | -2.2 | -1.7 | -1.0 | -.9 | -1.5 | 2.4 | -1.6 |
| | df | 843.8 | . | 155.2 | 158.7 | 59.9 | 195.9 | 192.2 | 181.9 | 165.6 | 169.0 | 159.2 | 156.0 | 243.5 | 56.3 | 67.3 | 165.7 | 56.3 |
| | P(2-tail) | .11 | . | .026 | .016 | .36 | .21 | .55 | .00 | .07 | .00 | .03 | .08 | .31 | .39 | .13 | .016 | .11 |
| | # Present | 950 | 950 | 934 | 932 | 664 | 907 | 933 | 923 | 919 | 916 | 917 | 917 | 928 | 744 | 783 | 851 | 767 |
| | # Missing | 440 | 0 | 126 | 126 | 51 | 167 | 163 | 144 | 137 | 137 | 134 | 135 | 185 | 53 | 66 | 134 | 57 |
| | Mean(Present) | 73.6 | 28.6 | 137.4 | 77.7 | 73.0 | 60.3 | 7.4 | 4.4 | 1.2 | 2.5 | 3.7 | 1.6 | 94.4 | 8.3 | 40.0 | 66.1 | 5.6 |
| | Mean(Missing) | 72.6 | . | 141.6 | 80.3 | 74.5 | 70.8 | 7.5 | 4.9 | 1.3 | 2.9 | 4.0 | 1.8 | 98.6 | 9.1 | 86.1 | 61.3 | 25.6 |
| SBP | t | 1.5 | -1.3 | . | . | 8.7 | -1.9 | -.6 | -3.2 | -.3 | -3.3 | -1.9 | -.5 | .3 | -1.6 | -.6 | 1.5 | -.6 |
| | df | 569.4 | 15.2 | . | . | 1.2 | 90.6 | 93.5 | 88.2 | 85.3 | 83.0 | 80.6 | 92.4 | 134.6 | 26.9 | 34.7 | 74.7 | 26.0 |
| | P(2-tail) | .135 | .204 | . | . | .05 | .06 | .53 | .00 | .76 | .00 | .06 | .62 | .78 | .11 | .56 | .14 | .57 |
| | # Present | 1060 | 934 | 1060 | 1058 | 713 | 986 | 1011 | 989 | 981 | 979 | 978 | 978 | 1013 | 770 | 814 | 919 | 798 |
| | # Missing | 330 | 16 | 0 | 0 | 2 | 88 | 85 | 78 | 75 | 74 | 73 | 74 | 100 | 27 | 35 | 66 | 26 |
| | Mean(Present) | 73.5 | 28.5 | 137.9 | 78.0 | 73.1 | 59.5 | 7.4 | 4.4 | 1.2 | 2.5 | 3.7 | 1.6 | 95.2 | 8.3 | 42.6 | 65.6 | 6.8 |
| | Mean(Missing) | 72.5 | 31.2 | . | . | 59.5 | 88.9 | 7.6 | 4.9 | 1.2 | 3.0 | 4.1 | 1.7 | 94.0 | 10.5 | 67.1 | 62.1 | 11.1 |
| DBP | t | 1.5 | -1.1 | 32.0 | . | 8.7 | -1.9 | -.6 | -3.5 | -.4 | -3.6 | -2.0 | -.6 | .3 | -1.6 | -.5 | 1.5 | -.5 |
| | df | 576.2 | 17.3 | 1.8 | . | 1.2 | 92.9 | 96.2 | 90.7 | 88.1 | 85.6 | 83.2 | 95.7 | 139.1 | 28.0 | 35.8 | 77.6 | 27.1 |
| | P(2-tail) | .13 | .28 | .00 | . | .05 | .06 | .55 | .00 | .70 | .00 | .05 | .57 | .77 | .13 | .58 | .13 | .60 |
| | # Present | 1058 | 932 | 1058 | 1058 | 713 | 984 | 1009 | 987 | 979 | 977 | 976 | 976 | 1011 | 769 | 813 | 917 | 797 |
| | # Missing | 332 | 18 | 2 | 0 | 2 | 90 | 87 | 80 | 77 | 76 | 75 | 76 | 102 | 28 | 36 | 68 | 27 |
| | Mean(Present) | 73.5 | 28.5 | 137.9 | 78.0 | 73.1 | 59.5 | 7.4 | 4.4 | 1.2 | 2.5 | 3.7 | 1.6 | 95.2 | 8.3 | 42.6 | 65.7 | 6.8 |
| | Mean(Missing) | 72.60 | 30.5641 | 101.0000 | . | 59.5000 | 88.0392 | 7.6356 | 4.9691 | 1.2775 | 3.0239 | 4.1213 | 1.7558 | 94.0784 | 10.4000 | 65.2722 | 62.1176 | 10.7407 |
| HR | t | 3.0 | -.9 | .6 | -.5 | . | -3.6 | -.8 | -2.5 | -.5 | -1.5 | -1.6 | -1.7 | 1.5 | -.1 | .4 | 2.2 | .9 |
| | df | 1380.3 | 524.2 | 652.5 | 722.2 | . | 465.2 | 765.2 | 805.8 | 735.4 | 758.9 | 742.6 | 820.6 | 991.6 | 340.0 | 397.6 | 710.6 | 654.6 |
| | P(2-tail) | .00 | .35 | .58 | .59 | . | .00 | .44 | .01 | .63 | .13 | .10 | .09 | .12 | .94 | .66 | .03 | .38 |
| | # Present | 715 | 664 | 713 | 713 | 715 | 674 | 692 | 675 | 675 | 677 | 672 | 675 | 693 | 593 | 601 | 673 | 602 |
| | # Missing | 675 | 286 | 347 | 345 | 0 | 400 | 404 | 392 | 381 | 376 | 379 | 377 | 420 | 204 | 248 | 312 | 222 |
| | Mean(Present) | 74.1 | 28.5 | 138.1 | 77.9 | 73.1 | 54.0 | 7.4 | 4.4 | 1.2 | 2.5 | 3.7 | 1.6 | 96.7 | 8.4 | 44.9 | 66.3 | 7.4 |
| | Mean(Missing) | 72.4 | 28.8 | 137.4 | 78.3 | . | 75.3 | 7.5 | 4.6 | 1.2 | 2.6 | 3.8 | 1.7 | 92.3 | 8.4 | 40.4 | 63.6 | 5.7 |
| HbA1c | t | .9 | .6 | -.9 | -1.4 | -1.4 | . | 4.9 | -4.5 | -1.2 | -3.7 | -1.6 | -.4 | -.9 | -.4 | 2.2 | 1.1 | 3.4 |
| | df | 548.0 | 46.7 | 84.3 | 95.1 | 45.4 | . | 56.2 | 42.8 | 40.5 | 40.9 | 40.7 | 38.8 | 79.3 | 13.4 | 22.7 | 72.1 | 61.5 |
| | P(2-tail) | .361 | .573 | .345 | .165 | .177 | . | .000 | .000 | .225 | .001 | .117 | .690 | .365 | .696 | .038 | .266 | .001 |
| | # Present | 1074 | 907 | 986 | 984 | 674 | 1074 | 1049 | 1027 | 1017 | 1014 | 1012 | 1015 | 1042 | 783 | 832 | 921 | 809 |
| | # Missing | 316 | 43 | 74 | 74 | 41 | 0 | 47 | 40 | 39 | 39 | 39 | 37 | 71 | 14 | 17 | 64 | 15 |
| | Mean(Present) | 73.4 | 28.6 | 137.7 | 77.9 | 72.9 | 61.9 | 7.5 | 4.5 | 1.2 | 2.6 | 3.7 | 1.6 | 94.7 | 8.4 | 44.1 | 65.6 | 7.0 |
| | Mean(Missing) | 72.8 | 28.2 | 139.9 | 79.3 | 75.6 | . | 6.3 | 5.2 | 1.3 | 3.1 | 4.1 | 1.7 | 100.2 | 8.9 | 19.4 | 62.8 | 2.0 |
| FG | t | .7 | .4 | -.3 | .4 | -1.0 | 1.9 | . | -.7 | -.7 | .2 | .3 | .6 | -1.9 | -.8 | 5.4 | 2.1 | 5.9 |
| | df | 488.4 | 16.5 | 51.3 | 55.3 | 23.6 | 71.6 | . | 13.2 | 13.1 | 12.2 | 11.3 | 10.6 | 50.0 | 5.0 | 19.6 | 42.8 | 559.5 |
| | P(2-tail) | .461 | .694 | .776 | .712 | .334 | .061 | . | .469 | .493 | .857 | .739 | .591 | .065 | .475 | .000 | .040 | .000 |
| | # Present | 1096 | 933 | 1011 | 1009 | 692 | 1049 | 1096 | 1053 | 1042 | 1040 | 1039 | 1041 | 1065 | 791 | 843 | 944 | 818 |
| | # Missing | 294 | 17 | 49 | 49 | 23 | 25 | 0 | 14 | 14 | 13 | 12 | 11 | 48 | 6 | 6 | 41 | 6 |

| | | | | | | | | | | | | | | | | | | |
|---|---|---|---|---|---|---|---|---|---|---|---|---|---|---|---|---|---|---|
| | Mean(Present) | 73.4 | 28.6 | 137.8 | 78.0 | 73.0 | 62.1 | 7.4 | 4.5 | 1.2 | 2.6 | 3.8 | 1.7 | 94.4 | 8.3 | 43.8 | 65.7 | 7.0 |
| | Mean(Missing) | 72.9 | 28.0 | 138.8 | 77.5 | 75.5 | 54.9 | . | 4.8 | 1.4 | 2.5 | 3.6 | 1.5 | 110.2 | 10.3 | 9.6 | 58.6 | .5 |
| TC | t | -.2 | .7 | -1.2 | -1.2 | -.7 | -.8 | -1.2 | . | . | 6.4 | . | . | -2.4 | .0 | 6.1 | 2.9 | 5.3 |
| | df | 548.6 | 27.3 | 78.1 | 84.2 | 42.3 | 47.3 | 43.0 | . | . | 1.1 | . | . | 69.8 | 4.0 | 9.7 | 62.9 | 138.4 |
| | P(2-tail) | .857 | .492 | .254 | .253 | .471 | .454 | .236 | . | . | .087 | . | . | .018 | .965 | .000 | .006 | .000 |
| | # Present | 1067 | 923 | 989 | 987 | 675 | 1027 | 1053 | 1067 | 1056 | 1051 | 1051 | 1052 | 1047 | 792 | 846 | 927 | 819 |
| | # Missing | 323 | 27 | 71 | 71 | 40 | 47 | 43 | 0 | 0 | 2 | 0 | 0 | 66 | 5 | 3 | 58 | 5 |
| | Mean(Present) | 73.2 | 28.6 | 137.7 | 77.9 | 73.0 | 61.3 | 7.4 | 4.5 | 1.2 | 2.6 | 3.8 | 1.6 | 93.9 | 8.4 | 43.7 | 65.9 | 7.0 |
| | Mean(Missing) | 73.4 | 27.8 | 140.6 | 79.3 | 74.7 | 76.0 | 8.2 | . | . | 1.6 | . | . | 112.8 | 8.3 | 7.3 | 57.8 | .9 |
| HDL | t | -.1 | .7 | -1.7 | -1.8 | -.7 | -.6 | -1.0 | -1.4 | . | 6.4 | 1.1 | . | -2.6 | -.4 | 6.3 | 3.2 | 5.9 |
| | df | 577.6 | 31.8 | 88.2 | 94.5 | 42.3 | 58.5 | 54.9 | 10.2 | . | 1.1 | 3.1 | . | 73.4 | 6.0 | 29.3 | 66.6 | 614.4 |
| | P(2-tail) | .891 | .487 | .093 | .077 | .471 | .541 | .307 | .188 | . | .087 | .362 | . | .012 | .727 | .000 | .002 | .000 |
| | # Present | 1056 | 919 | 981 | 979 | 675 | 1017 | 1042 | 1056 | 1056 | 1051 | 1047 | 1051 | 1044 | 790 | 843 | 924 | 815 |
| | # Missing | 334 | 31 | 79 | 79 | 40 | 57 | 54 | 11 | 0 | 2 | 4 | 1 | 69 | 7 | 6 | 61 | 9 |
| | Mean(Present) | 73.2 | 28.6 | 137.6 | 77.8 | 73.0 | 61.4 | 7.4 | 4.5 | 1.2 | 2.6 | 3.8 | 1.7 | 93.9 | 8.4 | 43.8 | 65.9 | 7.0 |
| | Mean(Missing) | 73.3 | 27.9 | 141.7 | 80.0 | 74.7 | 71.4 | 7.9 | 5.0 | . | 1.6 | 3.3 | 1.2 | 113.3 | 9.6 | 7.3 | 57.3 | .5 |
| LDL | t | -.1 | .1 | -1.9 | -1.8 | -.9 | -.6 | -1.3 | -1.4 | .2 | . | .3 | -.1 | -2.2 | -.6 | 6.3 | 2.7 | 5.9 |
| | df | 585.9 | 35.0 | 91.0 | 96.5 | 39.9 | 61.9 | 57.0 | 15.3 | 4.0 | . | 6.1 | 2.0 | 81.7 | 5.0 | 29.3 | 69.6 | 614.4 |
| | P(2-tail) | .940 | .904 | .059 | .078 | .378 | .542 | .209 | .185 | .879 | . | .795 | .913 | .029 | .595 | .000 | .008 | .000 |
| | # Present | 1053 | 916 | 979 | 977 | 677 | 1014 | 1040 | 1051 | 1051 | 1053 | 1044 | 1049 | 1040 | 791 | 843 | 922 | 815 |
| | # Missing | 337 | 34 | 81 | 81 | 38 | 60 | 56 | 16 | 5 | 0 | 7 | 3 | 73 | 6 | 6 | 63 | 9 |
| | Mean(Present) | 73.3 | 28.6 | 137.5 | 77.8 | 73.0 | 61.4 | 7.4 | 4.5 | 1.2 | 2.6 | 3.8 | 1.6 | 94.2 | 8.3 | 43.8 | 65.9 | 7.0 |
| | Mean(Missing) | 73.3 | 28.5 | 142.1 | 80.0 | 75.1 | 70.8 | 8.0 | 4.9 | 1.2 | . | 3.6 | 1.7 | 107.6 | 10.5 | 7.3 | 58.7 | .5 |
| TC/HDL | t | -.3 | .4 | -1.8 | -1.4 | -.7 | -.6 | -1.1 | -3.7 | -.7 | -1.0 | . | -.3 | -2.5 | -.4 | 6.3 | 3.3 | 5.8 |
| | df | 590.3 | 34.0 | 92.7 | 98.7 | 46.2 | 64.2 | 58.1 | 15.6 | 8.1 | 8.1 | . | 8.3 | 84.6 | 6.0 | 29.3 | 72.9 | 675.7 |
| | P(2-tail) | .773 | .721 | .082 | .173 | .473 | .572 | .296 | .002 | .530 | .364 | . | .773 | .013 | .727 | .000 | .002 | .000 |
| | # Present | 1051 | 917 | 978 | 976 | 672 | 1012 | 1039 | 1051 | 1047 | 1044 | 1051 | 1043 | 1035 | 790 | 843 | 919 | 814 |
| | # Missing | 339 | 33 | 82 | 82 | 43 | 62 | 57 | 16 | 9 | 9 | 0 | 9 | 78 | 7 | 6 | 66 | 10 |
| | Mean(Present) | 73.2 | 28.6 | 137.6 | 77.9 | 73.0 | 61.4 | 7.4 | 4.5 | 1.2 | 2.6 | 3.8 | 1.6 | 93.9 | 8.4 | 43.8 | 66.0 | 7.0 |
| | Mean(Missing) | 73.4 | 28.2 | 141.7 | 79.5 | 74.6 | 69.9 | 7.9 | 5.4 | 1.3 | 2.9 | . | 1.7 | 111.2 | 9.6 | 7.3 | 57.4 | .6 |
| TG | t | -.1 | .2 | -1.9 | -1.8 | -.7 | -.6 | -.9 | -1.5 | .2 | 1.2 | -.1 | . | -2.4 | -.8 | 5.3 | 3.0 | 5.8 |
| | df | 592.4 | 33.9 | 92.2 | 98.2 | 42.3 | 60.7 | 56.0 | 14.3 | 4.0 | 3.0 | 7.1 | . | 78.4 | 7.0 | 23.3 | 70.5 | 662.9 |
| | P(2-tail) | .927 | .804 | .056 | .076 | .471 | .562 | .356 | .166 | .875 | .318 | .953 | . | .018 | .470 | .000 | .004 | .000 |
| | # Present | 1052 | 917 | 978 | 976 | 675 | 1015 | 1041 | 1052 | 1051 | 1049 | 1043 | 1052 | 1040 | 789 | 842 | 921 | 814 |
| | # Missing | 338 | 33 | 82 | 82 | 40 | 59 | 55 | 15 | 5 | 4 | 8 | 0 | 73 | 8 | 7 | 64 | 10 |
| | Mean(Present) | 73.3 | 28.6 | 137.5 | 77.8 | 73.0 | 61.4 | 7.4 | 4.5 | 1.2 | 2.6 | 3.8 | 1.6 | 93.9 | 8.3 | 43.9 | 65.9 | 7.0 |
| | Mean(Missing) | 73.3 | 28.3 | 142.1 | 80.0 | 74.7 | 70.6 | 7.9 | 5.0 | 1.2 | 2.0 | 3.8 | . | 111.3 | 10.8 | 10.5 | 58.0 | .6 |
| Creat B | t | 1.1 | .0 | -.7 | -2.0 | .7 | -.8 | .1 | -.9 | -.9 | .0 | 2.0 | 2.1 | . | .1 | 3.2 | . | 3.9 |
| | df | 462.4 | 22.0 | 48.7 | 51.3 | 22.4 | 31.4 | 31.1 | 19.7 | 11.2 | 12.2 | 16.3 | 11.8 | . | 2.0 | 4.8 | . | 16.2 |
| | P(2-tail) | .254 | .986 | .485 | .049 | .498 | .457 | .890 | .378 | .383 | .971 | .062 | .057 | . | .899 | .027 | . | .001 |
| | # Present | 1113 | 928 | 1013 | 1011 | 693 | 1042 | 1065 | 1047 | 1044 | 1040 | 1035 | 1040 | 1113 | 794 | 845 | 985 | 819 |
| | # Missing | 277 | 22 | 47 | 47 | 22 | 32 | 31 | 20 | 12 | 13 | 16 | 12 | 0 | 3 | 4 | 0 | 5 |
| | Mean(Present) | 73.4 | 28.6 | 137.8 | 77.9 | 73.1 | 61.3 | 7.4 | 4.5 | 1.2 | 2.6 | 3.8 | 1.7 | 95.1 | 8.4 | 43.7 | 65.4 | 7.0 |
| | Mean(Missing) | 72.7 | 28.6 | 140.2 | 80.9 | 71.3 | 82.7 | 7.3 | 4.7 | 1.3 | 2.6 | 3.4 | 1.3 | . | 7.7 | 12.7 | . | 1.2 |

| | | | | | | | | | | | | | | | | | | |
|---|---|---|---|---|---|---|---|---|---|---|---|---|---|---|---|---|---|---|
| Creat U | t | 2.3 | .3 | .1 | -1.6 | -.4 | -3.5 | 1.1 | -5.3 | -1.9 | -3.8 | -2.0 | -.9 | .3 | . | 3.2 | 5.2 | 3.1 |
| | df | 1209.3 | 324.2 | 531.7 | 554.0 | 186.2 | 320.8 | 515.1 | 447.0 | 413.0 | 411.9 | 410.9 | 496.6 | 577.7 | . | 320.1 | 417.1 | 366.1 |
| | P(2-tail) | .019 | .759 | .952 | .115 | .702 | .001 | .258 | .000 | .065 | .000 | .046 | .351 | .730 | . | .001 | .000 | .002 |
| | # Present | 797 | 744 | 770 | 769 | 593 | 783 | 791 | 792 | 790 | 791 | 790 | 789 | 794 | 797 | 754 | 760 | 775 |
| | # Missing | 593 | 206 | 290 | 289 | 122 | 291 | 305 | 275 | 266 | 262 | 261 | 263 | 319 | 0 | 95 | 225 | 49 |
| | Mean(Present) | 73.8 | 28.6 | 137.9 | 77.7 | 73.0 | 54.7 | 7.5 | 4.4 | 1.2 | 2.5 | 3.7 | 1.6 | 95.4 | 8.4 | 46.1 | 67.0 | 7.2 |
| | Mean(Missing) | 72.5 | 28.5 | 137.8 | 78.8 | 73.5 | 81.4 | 7.3 | 4.8 | 1.3 | 2.8 | 3.9 | 1.7 | 94.3 | . | 23.4 | 60.1 | 3.0 |
| Alb U | t | -.5 | .4 | .1 | -.4 | -1.0 | 1.0 | 1.0 | -4.0 | .6 | -3.6 | -3.4 | -1.6 | .4 | -1.1 | . | 3.3 | 3.8 |
| | df | 1195.2 | 261.5 | 422.5 | 450.4 | 163.0 | 441.4 | 375.6 | 313.0 | 316.2 | 292.8 | 280.1 | 308.2 | 535.0 | 45.3 | . | 340.5 | 727.8 |
| | P(2-tail) | .598 | .722 | .911 | .684 | .332 | .336 | .339 | .000 | .528 | .000 | .001 | .121 | .721 | .280 | . | .001 | .000 |
| | # Present | 849 | 783 | 814 | 813 | 601 | 832 | 843 | 846 | 843 | 843 | 843 | 842 | 845 | 754 | 849 | 784 | 757 |
| | # Missing | 541 | 167 | 246 | 245 | 114 | 242 | 253 | 221 | 213 | 210 | 208 | 210 | 268 | 43 | 0 | 201 | 67 |
| | Mean(Present) | 73.2 | 28.6 | 137.9 | 77.9 | 72.9 | 63.1 | 7.5 | 4.4 | 1.2 | 2.5 | 3.7 | 1.6 | 95.3 | 8.3 | 43.6 | 66.3 | 7.3 |
| | Mean(Missing) | 73.4 | 28.5 | 137.8 | 78.2 | 74.1 | 57.9 | 7.3 | 4.8 | 1.2 | 2.8 | 4.1 | 1.8 | 94.2 | 9.3 | . | 61.8 | 2.5 |
| eGFR | t | 4.0 | -1.4 | -.5 | -3.7 | .2 | -2.9 | -.6 | -3.3 | -3.4 | -.7 | 1.8 | .1 | 4.8 | .5 | .0 | . | -.5 |
| | df | 768.2 | 118.5 | 175.0 | 196.5 | 47.6 | 160.4 | 197.9 | 190.2 | 166.5 | 173.8 | 183.5 | 188.4 | 278.7 | 38.5 | 69.9 | . | 19.8 |
| | P(2-tail) | .000 | .165 | .638 | .000 | .861 | .004 | .567 | .001 | .001 | .472 | .076 | .949 | .000 | .651 | .981 | . | .626 |
| | # Present | 985 | 851 | 919 | 917 | 673 | 921 | 944 | 927 | 924 | 922 | 919 | 921 | 985 | 760 | 784 | 985 | 804 |
| | # Missing | 405 | 99 | 141 | 141 | 42 | 153 | 152 | 140 | 132 | 131 | 132 | 131 | 128 | 37 | 65 | 0 | 20 |
| | Mean(Present) | 74.0 | 28.5 | 137.8 | 77.5 | 73.1 | 56.9 | 7.4 | 4.4 | 1.2 | 2.6 | 3.8 | 1.7 | 96.6 | 8.4 | 43.6 | 65.4 | 6.9 |
| | Mean(Missing) | 71.6 | 29.3 | 138.6 | 81.0 | 72.8 | 91.8 | 7.5 | 4.8 | 1.3 | 2.6 | 3.6 | 1.6 | 83.3 | 7.9 | 43.1 | . | 10.5 |
| UACR | t | 2.6 | .0 | -.9 | -2.4 | -1.0 | -3.1 | .7 | -5.4 | -2.7 | -3.9 | -1.2 | -.2 | -.1 | -.3 | 3.5 | 4.4 | . |
| | df | 1159.5 | 273.7 | 437.0 | 463.7 | 162.8 | 293.8 | 432.5 | 388.9 | 358.7 | 368.7 | 366.6 | 447.2 | 464.2 | 21.6 | 325.4 | 272.6 | . |
| | P(2-tail) | .008 | .965 | .360 | .016 | .332 | .002 | .480 | .000 | .007 | .000 | .213 | .807 | .906 | .734 | .001 | .000 | . |
| | # Present | 824 | 767 | 798 | 797 | 602 | 809 | 818 | 819 | 815 | 815 | 814 | 814 | 819 | 775 | 757 | 804 | 824 |
| | # Missing | 566 | 183 | 262 | 261 | 113 | 265 | 278 | 248 | 241 | 238 | 237 | 238 | 294 | 22 | 92 | 181 | 0 |
| | Mean(Present) | 73.9 | 28.6 | 137.6 | 77.5 | 72.9 | 55.7 | 7.4 | 4.4 | 1.2 | 2.5 | 3.7 | 1.6 | 95.0 | 8.3 | 46.2 | 66.7 | 7.0 |
| | Mean(Missing) | 72.4 | 28.6 | 138.8 | 79.4 | 74.1 | 80.7 | 7.3 | 4.8 | 1.3 | 2.8 | 3.9 | 1.7 | 95.4 | 8.8 | 22.0 | 59.9 | . |
| Smoking behavior | t | 1.5 | -.8 | -2.4 | -2.2 | -1.1 | -2.9 | -1.2 | -3.0 | -3.0 | -1.0 | .8 | -.5 | -2.0 | -.9 | -1.0 | 4.2 | -1.5 |
| | df | 1165.0 | 197.0 | 371.9 | 412.4 | 129.8 | 324.8 | 378.0 | 410.6 | 354.1 | 389.7 | 415.1 | 364.6 | 503.6 | 148.3 | 206.9 | 292.7 | 116.7 |
| | P(2-tail) | .144 | .440 | .015 | .029 | .279 | .004 | .242 | .002 | .003 | .314 | .439 | .608 | .051 | .386 | .322 | .000 | .148 |
| | # Present | 836 | 805 | 815 | 814 | 609 | 800 | 818 | 813 | 811 | 810 | 808 | 810 | 820 | 680 | 685 | 787 | 709 |
| | # Missing | 554 | 145 | 245 | 244 | 106 | 274 | 278 | 254 | 245 | 243 | 243 | 242 | 293 | 117 | 164 | 198 | 115 |
| | Mean(Present) | 73.6 | 28.5 | 137.1 | 77.6 | 72.8 | 56.4 | 7.4 | 4.4 | 1.2 | 2.6 | 3.8 | 1.6 | 93.4 | 8.3 | 41.0 | 66.7 | 5.7 |
| | Mean(Missing) | 72.8 | 28.9 | 140.6 | 79.3 | 74.5 | 77.8 | 7.6 | 4.7 | 1.3 | 2.6 | 3.7 | 1.7 | 99.8 | 8.8 | 54.4 | 60.2 | 14.8 |
| eGFR categories | t | 4.0 | -1.4 | -.5 | -3.7 | .2 | -2.9 | -.6 | -3.3 | -3.4 | -.7 | 1.8 | .1 | 4.8 | .5 | .0 | . | -.5 |
| | df | 768.2 | 118.5 | 175.0 | 196.5 | 47.6 | 160.4 | 197.9 | 190.2 | 166.5 | 173.8 | 183.5 | 188.4 | 278.7 | 38.5 | 69.9 | . | 19.8 |
| | P(2-tail) | .000 | .165 | .638 | .000 | .861 | .004 | .567 | .001 | .001 | .472 | .076 | .949 | .000 | .651 | .981 | . | .626 |
| | # Present | 985 | 851 | 919 | 917 | 673 | 921 | 944 | 927 | 924 | 922 | 919 | 921 | 985 | 760 | 784 | 985 | 804 |
| | # Missing | 405 | 99 | 141 | 141 | 42 | 153 | 152 | 140 | 132 | 131 | 132 | 131 | 128 | 37 | 65 | 0 | 20 |
| | Mean(Present) | 74.0 | 28.5 | 137.8 | 77.5 | 73.1 | 56.9 | 7.4 | 4.4 | 1.2 | 2.6 | 3.8 | 1.7 | 96.6 | 8.4 | 43.6 | 65.4 | 6.9 |
| | Mean(Missing) | 71.6 | 29.3 | 138.6 | 81.0 | 72.8 | 91.8 | 7.5 | 4.8 | 1.3 | 2.6 | 3.6 | 1.6 | 83.3 | 7.9 | 43.1 | . | 10.5 |
| Albuminuria | t | 2.6 | .0 | -.9 | -2.4 | -1.0 | -3.1 | .7 | -5.4 | -2.7 | -3.9 | -1.2 | -.2 | -.1 | -.3 | 3.5 | 4.4 | . |
| | df | 1159.5 | 273.7 | 437.0 | 463.7 | 162.8 | 293.8 | 432.5 | 388.9 | 358.7 | 368.7 | 366.6 | 447.2 | 464.2 | 21.6 | 325.4 | 272.6 | . |

| | | | | | | | | | | | | | | | | | |
|---|---|---|---|---|---|---|---|---|---|---|---|---|---|---|---|---|---|
| P(2-tail) | .008 | .965 | .360 | .016 | .332 | .002 | .480 | .000 | .007 | .000 | .213 | .807 | .906 | .734 | .001 | .000 | . |
| # Present | 824 | 767 | 798 | 797 | 602 | 809 | 818 | 819 | 815 | 815 | 814 | 814 | 819 | 775 | 757 | 804 | 824 |
| # Missing | 566 | 183 | 262 | 261 | 113 | 265 | 278 | 248 | 241 | 238 | 237 | 238 | 294 | 22 | 92 | 181 | 0 |
| Mean(Present) | 73.9 | 28.6 | 137.6 | 77.5 | 72.9 | 55.7 | 7.4 | 4.4 | 1.2 | 2.5 | 3.7 | 1.6 | 95.0 | 8.3 | 46.2 | 66.7 | 7.0 |
| Mean(Missing) | 72.4 | 28.6 | 138.8 | 79.4 | 74.1 | 80.7 | 7.3 | 4.8 | 1.3 | 2.8 | 3.9 | 1.7 | 95.4 | 8.8 | 22.0 | 59.9 | . |

BMI, Body mass index; SBP, Systolic blood pressure; DBP, Diastolic blood pressure; HR, Heart rate; HbA1c, Glycated haemoglobin A1c; FG, FG; TC, TC; HDL, high density lipoprotein cholesterol; LDL, low density lipoprotein cholesterol; TC/HDL, Total/HDL cholesterol ratio; TG, TG; Creat B, Creat B; Creat U, Creat U; Alb U, Albumin urine; eGFR, estimated Glomerular filtration rate; UACR, Urinary albumin creatinine ratio.

For each quantitative variable, pairs of groups are formed by indicator variables (present, missing).
a. Indicator variables with less than 5% missing are not displayed.

#### Tables S3.2-S3.12. Cross tabulations of categorical versus missingness indicator variables

**Table S3.2**

| Sex | | | Total | Female | Male |
|---|---|---|---|---|---|
| BMI | Present | N | 950 | 322 | 628 |
| | | % | 68.3 | 65.1 | 70.2 |
| | Missing | % | 31.7 | 34.9 | 29.8 |
| SBP | Present | N | 1060 | 373 | 687 |
| | | % | 76.3 | 75.4 | 76.8 |
| | Missing | % | 23.7 | 24.6 | 23.2 |
| DBP | Present | N | 1058 | 372 | 686 |
| | | % | 76.1 | 75.2 | 76.6 |
| | Missing | % | 23.9 | 24.8 | 23.4 |
| HR | Present | N | 715 | 242 | 473 |
| | | % | 51.4 | 48.9 | 52.8 |
| | Missing | % | 48.6 | 51.1 | 47.2 |
| HbA1c | Present | N | 1074 | 379 | 695 |
| | | % | 77.3 | 76.6 | 77.7 |
| | Missing | % | 22.7 | 23.4 | 22.3 |
| FG | Present | N | 1096 | 390 | 706 |
| | | % | 78.8 | 78.8 | 78.9 |
| | Missing | % | 21.2 | 21.2 | 21.1 |
| TC | Present | N | 1067 | 375 | 692 |
| | | % | 76.8 | 75.8 | 77.3 |
| | Missing | % | 23.2 | 24.2 | 22.7 |
| HDL | Present | N | 1056 | 374 | 682 |
| | | % | 76.0 | 75.6 | 76.2 |
| | Missing | % | 24.0 | 24.4 | 23.8 |
| LDL | Present | N | 1053 | 372 | 681 |
| | | % | 75.8 | 75.2 | 76.1 |
| | Missing | % | 24.2 | 24.8 | 23.9 |
| TC/HDL | Present | N | 1051 | 369 | 682 |
| | | % | 75.6 | 74.5 | 76.2 |
| | Missing | % | 24.4 | 25.5 | 23.8 |
| TG | Present | N | 1052 | 371 | 681 |
| | | % | 75.7 | 74.9 | 76.1 |
| | Missing | % | 24.3 | 25.1 | 23.9 |
| Creat B | Present | N | 1113 | 397 | 716 |
| | | % | 80.1 | 80.2 | 80.0 |
| | Missing | % | 19.9 | 19.8 | 20.0 |
| Creat U | Present | N | 797 | 268 | 529 |
| | | % | 57.3 | 54.1 | 59.1 |
| | Missing | % | 42.7 | 45.9 | 40.9 |
| Alb U | Present | N | 849 | 300 | 549 |
| | | % | 61.1 | 60.6 | 61.3 |
| | Missing | % | 38.9 | 39.4 | 38.7 |
| eGFR | Present | N | 985 | 336 | 649 |
| | | % | 70.9 | 67.9 | 72.5 |
| | Missing | % | 29.1 | 32.1 | 27.5 |
| UACR | Present | N | 824 | 277 | 547 |
| | | % | 59.3 | 56.0 | 61.1 |
| | Missing | % | 40.7 | 44.0 | 38.9 |
| Smoking Behaviour | Present | N | 836 | 286 | 550 |
| | | % | 60.1 | 57.8 | 61.5 |

| | | | | | |
|---|---|---|---|---|---|
| | Missing | % | 39.9 | 42.2 | 38.5 |
| eGFR categories | Present | N | 985 | 336 | 649 |
| | | % | 70.9 | 67.9 | 72.5 |
| | Missing | % | 29.1 | 32.1 | 27.5 |
| Albuminuria | Present | N | 824 | 277 | 547 |
| | | % | 59.3 | 56.0 | 61.1 |
| | Missing | % | 40.7 | 44.0 | 38.9 |

Indicator variables with less than 5% missing are not displayed.

**Table S3.3**

| eGFR categories | | | Total | normal or high (>90 ml/min) | mildly decreased (60-90 ml/min) | moderately decreased (30-60 ml/min) | severely decreased (<30 ml/min) | Missing |
|---|---|---|---|---|---|---|---|---|
| BMI | Present | N | 950 | 103 | 511 | 206 | 31 | 99 |
| | | % | 68.3 | 86.6 | 88.9 | 83.1 | 72.1 | 24.4 |
| | Missing | % | 31.7 | 13.4 | 11.1 | 16.9 | 27.9 | 75.6 |
| SBP | Present | N | 1060 | 113 | 539 | 228 | 39 | 141 |
| | | % | 76.3 | 95.0 | 93.7 | 91.9 | 90.7 | 34.8 |
| | Missing | % | 23.7 | 5.0 | 6.3 | 8.1 | 9.3 | 65.2 |
| DBP | Present | N | 1058 | 113 | 538 | 227 | 39 | 141 |
| | | % | 76.1 | 95.0 | 93.6 | 91.5 | 90.7 | 34.8 |
| | Missing | % | 23.9 | 5.0 | 6.4 | 8.5 | 9.3 | 65.2 |
| HR | Present | N | 715 | 93 | 385 | 162 | 33 | 42 |
| | | % | 51.4 | 78.2 | 67.0 | 65.3 | 76.7 | 10.4 |
| | Missing | % | 48.6 | 21.8 | 33.0 | 34.7 | 23.3 | 89.6 |
| HbA1c | Present | N | 1074 | 113 | 536 | 233 | 39 | 153 |
| | | % | 77.3 | 95.0 | 93.2 | 94.0 | 90.7 | 37.8 |
| | Missing | % | 22.7 | 5.0 | 6.8 | 6.0 | 9.3 | 62.2 |
| FG | Present | N | 1096 | 115 | 557 | 233 | 39 | 152 |
| | | % | 78.8 | 96.6 | 96.9 | 94.0 | 90.7 | 37.5 |
| | Missing | % | 21.2 | 3.4 | 3.1 | 6.0 | 9.3 | 62.5 |
| TC | Present | N | 1067 | 114 | 550 | 226 | 37 | 140 |
| | | % | 76.8 | 95.8 | 95.7 | 91.1 | 86.0 | 34.6 |
| | Missing | % | 23.2 | 4.2 | 4.3 | 8.9 | 14.0 | 65.4 |
| HDL | Present | N | 1056 | 114 | 549 | 225 | 36 | 132 |
| | | % | 76.0 | 95.8 | 95.5 | 90.7 | 83.7 | 32.6 |
| | Missing | % | 24.0 | 4.2 | 4.5 | 9.3 | 16.3 | 67.4 |
| LDL | Present | N | 1053 | 114 | 547 | 224 | 37 | 131 |
| | | % | 75.8 | 95.8 | 95.1 | 90.3 | 86.0 | 32.3 |
| | Missing | % | 24.2 | 4.2 | 4.9 | 9.7 | 14.0 | 67.7 |
| TC/HDL | Present | N | 1051 | 114 | 546 | 224 | 35 | 132 |
| | | % | 75.6 | 95.8 | 95.0 | 90.3 | 81.4 | 32.6 |
| | Missing | % | 24.4 | 4.2 | 5.0 | 9.7 | 18.6 | 67.4 |
| TG | Present | N | 1052 | 114 | 547 | 223 | 37 | 131 |
| | | % | 75.7 | 95.8 | 95.1 | 89.9 | 86.0 | 32.3 |
| | Missing | % | 24.3 | 4.2 | 4.9 | 10.1 | 14.0 | 67.7 |
| Creat B | Present | N | 1113 | 119 | 575 | 248 | 43 | 128 |
| | | % | 80.1 | 100.0 | 100.0 | 100.0 | 100.0 | 31.6 |
| | Missing | % | 19.9 | .0 | .0 | .0 | .0 | 68.4 |
| Creat U | Present | N | 797 | 110 | 429 | 190 | 31 | 37 |
| | | % | 57.3 | 92.4 | 74.6 | 76.6 | 72.1 | 9.1 |
| | Missing | % | 42.7 | 7.6 | 25.4 | 23.4 | 27.9 | 90.9 |
| Alb U | Present | N | 849 | 108 | 446 | 197 | 33 | 65 |

| | | | | | | | | |
|---|---|---|---|---|---|---|---|---|
| | | % | 61.1 | 90.8 | 77.6 | 79.4 | 76.7 | 16.0 |
| | Missing | % | 38.9 | 9.2 | 22.4 | 20.6 | 23.3 | 84.0 |
| eGFR | Present | N | 985 | 119 | 575 | 248 | 43 | 0 |
| | | % | 70.9 | 100.0 | 100.0 | 100.0 | 100.0 | .0 |
| | Missing | % | 29.1 | .0 | .0 | .0 | .0 | 100.0 |
| UACR | Present | N | 824 | 110 | 464 | 199 | 31 | 20 |
| | | % | 59.3 | 92.4 | 80.7 | 80.2 | 72.1 | 4.9 |
| | Missing | % | 40.7 | 7.6 | 19.3 | 19.8 | 27.9 | 95.1 |
| Smoking Behaviour | Present | N | 836 | 104 | 474 | 183 | 26 | 49 |
| | | % | 60.1 | 87.4 | 82.4 | 73.8 | 60.5 | 12.1 |
| | Missing | % | 39.9 | 12.6 | 17.6 | 26.2 | 39.5 | 87.9 |
| Albuminuria | Present | N | 824 | 110 | 464 | 199 | 31 | 20 |
| | | % | 59.3 | 92.4 | 80.7 | 80.2 | 72.1 | 4.9 |
| | Missing | % | 40.7 | 7.6 | 19.3 | 19.8 | 27.9 | 95.1 |

Indicator variables with less than 5% missing are not displayed.

**Table S3.4**

| **Albuminuria** | | | Total | normal/mild (<3 mg/mmol) | moderate (3-30 mg/mmol) | severe (>30 mg/mmol) | Missing |
|---|---|---|---|---|---|---|---|
| BMI | Present | N | 950 | 560 | 174 | 33 | 183 |
| | | % | 68.3 | 93.3 | 94.6 | 82.5 | 32.3 |
| | Missing | % | 31.7 | 6.7 | 5.4 | 17.5 | 67.7 |
| SBP | Present | N | 1060 | 581 | 179 | 38 | 262 |
| | | % | 76.3 | 96.8 | 97.3 | 95.0 | 46.3 |
| | Missing | % | 23.7 | 3.2 | 2.7 | 5.0 | 53.7 |
| DBP | Present | N | 1058 | 580 | 179 | 38 | 261 |
| | | % | 76.1 | 96.7 | 97.3 | 95.0 | 46.1 |
| | Missing | % | 23.9 | 3.3 | 2.7 | 5.0 | 53.9 |
| HR | Present | N | 715 | 439 | 131 | 32 | 113 |
| | | % | 51.4 | 73.2 | 71.2 | 80.0 | 20.0 |
| | Missing | % | 48.6 | 26.8 | 28.8 | 20.0 | 80.0 |
| HbA1c | Present | N | 1074 | 587 | 182 | 40 | 265 |
| | | % | 77.3 | 97.8 | 98.9 | 100.0 | 46.8 |
| | Missing | % | 22.7 | 2.2 | 1.1 | .0 | 53.2 |
| FG | Present | N | 1096 | 594 | 184 | 40 | 278 |
| | | % | 78.8 | 99.0 | 100.0 | 100.0 | 49.1 |
| | Missing | % | 21.2 | 1.0 | .0 | .0 | 50.9 |
| TC | Present | N | 1067 | 595 | 184 | 40 | 248 |
| | | % | 76.8 | 99.2 | 100.0 | 100.0 | 43.8 |
| | Missing | % | 23.2 | .8 | .0 | .0 | 56.2 |
| HDL | Present | N | 1056 | 591 | 184 | 40 | 241 |
| | | % | 76.0 | 98.5 | 100.0 | 100.0 | 42.6 |
| | Missing | % | 24.0 | 1.5 | .0 | .0 | 57.4 |

| | | | | | | | |
|---|---|---|---|---|---|---|---|
| LDL | Present | N | 1053 | 591 | 184 | 40 | 238 |
| | | % | 75.8 | 98.5 | 100.0 | 100.0 | 42.0 |
| | Missing | % | 24.2 | 1.5 | .0 | .0 | 58.0 |
| TC/HDL | Present | N | 1051 | 590 | 184 | 40 | 237 |
| | | % | 75.6 | 98.3 | 100.0 | 100.0 | 41.9 |
| | Missing | % | 24.4 | 1.7 | .0 | .0 | 58.1 |
| TG | Present | N | 1052 | 590 | 184 | 40 | 238 |
| | | % | 75.7 | 98.3 | 100.0 | 100.0 | 42.0 |
| | Missing | % | 24.3 | 1.7 | .0 | .0 | 58.0 |
| Creat B | Present | N | 1113 | 596 | 183 | 40 | 294 |
| | | % | 80.1 | 99.3 | 99.5 | 100.0 | 51.9 |
| | Missing | % | 19.9 | .7 | .5 | .0 | 48.1 |
| Creat U | Present | N | 797 | 564 | 171 | 40 | 22 |
| | | % | 57.3 | 94.0 | 92.9 | 100.0 | 3.9 |
| | Missing | % | 42.7 | 6.0 | 7.1 | .0 | 96.1 |
| Alb U | Present | N | 849 | 548 | 169 | 40 | 92 |
| | | % | 61.1 | 91.3 | 91.8 | 100.0 | 16.3 |
| | Missing | % | 38.9 | 8.7 | 8.2 | .0 | 83.7 |
| eGFR | Present | N | 985 | 587 | 178 | 39 | 181 |
| | | % | 70.9 | 97.8 | 96.7 | 97.5 | 32.0 |
| | Missing | % | 29.1 | 2.2 | 3.3 | 2.5 | 68.0 |
| UACR | Present | N | 824 | 600 | 184 | 40 | 0 |
| | | % | 59.3 | 100.0 | 100.0 | 100.0 | .0 |
| | Missing | % | 40.7 | .0 | .0 | .0 | 100.0 |
| Smoking Behaviour | Present | N | 836 | 515 | 163 | 31 | 127 |
| | | % | 60.1 | 85.8 | 88.6 | 77.5 | 22.4 |
| | Missing | % | 39.9 | 14.2 | 11.4 | 22.5 | 77.6 |
| eGFR categories | Present | N | 985 | 587 | 178 | 39 | 181 |
| | | % | 70.9 | 97.8 | 96.7 | 97.5 | 32.0 |
| | Missing | % | 29.1 | 2.2 | 3.3 | 2.5 | 68.0 |

Indicator variables with less than 5% missing are not displayed.

**Table S3.5**

| Glucose lowering medication | | | Total | no medication | oral only | insulin only | oral and insulin |
|---|---|---|---|---|---|---|---|
| BMI | Present | N | 950 | 204 | 580 | 23 | 143 |
| | | % | 68.3 | 47.3 | 81.7 | 44.2 | 72.6 |
| | Missing | % | 31.7 | 52.7 | 18.3 | 55.8 | 27.4 |
| SBP | Present | N | 1060 | 234 | 633 | 31 | 162 |
| | | % | 76.3 | 54.3 | 89.2 | 59.6 | 82.2 |
| | Missing | % | 23.7 | 45.7 | 10.8 | 40.4 | 17.8 |

| | | | | | | | |
|---|---|---|---|---|---|---|---|
| DBP | Present | N | 1058 | 232 | 633 | 31 | 162 |
| | | % | 76.1 | 53.8 | 89.2 | 59.6 | 82.2 |
| | Missing | % | 23.9 | 46.2 | 10.8 | 40.4 | 17.8 |
| HR | Present | N | 715 | 155 | 422 | 18 | 120 |
| | | % | 51.4 | 36.0 | 59.4 | 34.6 | 60.9 |
| | Missing | % | 48.6 | 64.0 | 40.6 | 65.4 | 39.1 |
| HbA1c | Present | N | 1074 | 215 | 663 | 32 | 164 |
| | | % | 77.3 | 49.9 | 93.4 | 61.5 | 83.2 |
| | Missing | % | 22.7 | 50.1 | 6.6 | 38.5 | 16.8 |
| FG | Present | N | 1096 | 248 | 658 | 30 | 160 |
| | | % | 78.8 | 57.5 | 92.7 | 57.7 | 81.2 |
| | Missing | % | 21.2 | 42.5 | 7.3 | 42.3 | 18.8 |
| TC | Present | N | 1067 | 240 | 642 | 27 | 158 |
| | | % | 76.8 | 55.7 | 90.4 | 51.9 | 80.2 |
| | Missing | % | 23.2 | 44.3 | 9.6 | 48.1 | 19.8 |
| HDL | Present | N | 1056 | 237 | 635 | 26 | 158 |
| | | % | 76.0 | 55.0 | 89.4 | 50.0 | 80.2 |
| | Missing | % | 24.0 | 45.0 | 10.6 | 50.0 | 19.8 |
| LDL | Present | N | 1053 | 236 | 635 | 26 | 156 |
| | | % | 75.8 | 54.8 | 89.4 | 50.0 | 79.2 |
| | Missing | % | 24.2 | 45.2 | 10.6 | 50.0 | 20.8 |
| TC/HDL | Present | N | 1051 | 234 | 634 | 26 | 157 |
| | | % | 75.6 | 54.3 | 89.3 | 50.0 | 79.7 |
| | Missing | % | 24.4 | 45.7 | 10.7 | 50.0 | 20.3 |
| TG | Present | N | 1052 | 235 | 634 | 26 | 157 |
| | | % | 75.7 | 54.5 | 89.3 | 50.0 | 79.7 |
| | Missing | % | 24.3 | 45.5 | 10.7 | 50.0 | 20.3 |
| Creat B | Present | N | 1113 | 251 | 651 | 37 | 174 |
| | | % | 80.1 | 58.2 | 91.7 | 71.2 | 88.3 |
| | Missing | % | 19.9 | 41.8 | 8.3 | 28.8 | 11.7 |
| Creat U | Present | N | 797 | 152 | 497 | 18 | 130 |
| | | % | 57.3 | 35.3 | 70.0 | 34.6 | 66.0 |
| | Missing | % | 42.7 | 64.7 | 30.0 | 65.4 | 34.0 |
| Alb U | Present | N | 849 | 161 | 529 | 21 | 138 |
| | | % | 61.1 | 37.4 | 74.5 | 40.4 | 70.1 |
| | Missing | % | 38.9 | 62.6 | 25.5 | 59.6 | 29.9 |
| eGFR | Present | N | 985 | 224 | 572 | 31 | 158 |
| | | % | 70.9 | 52.0 | 80.6 | 59.6 | 80.2 |
| | Missing | % | 29.1 | 48.0 | 19.4 | 40.4 | 19.8 |
| UACR | Present | N | 824 | 158 | 513 | 21 | 132 |

| | | | | | | | |
|---|---|---|---|---|---|---|---|
| | | % | 59.3 | 36.7 | 72.3 | 40.4 | 67.0 |
| | Missing | % | 40.7 | 63.3 | 27.7 | 59.6 | 33.0 |
| Smoking Behaviour | Present | N | 836 | 181 | 511 | 17 | 127 |
| | | % | 60.1 | 42.0 | 72.0 | 32.7 | 64.5 |
| | Missing | % | 39.9 | 58.0 | 28.0 | 67.3 | 35.5 |
| eGFR categories | Present | N | 985 | 224 | 572 | 31 | 158 |
| | | % | 70.9 | 52.0 | 80.6 | 59.6 | 80.2 |
| | Missing | % | 29.1 | 48.0 | 19.4 | 40.4 | 19.8 |
| Albuminuria | Present | N | 824 | 158 | 513 | 21 | 132 |
| | | % | 59.3 | 36.7 | 72.3 | 40.4 | 67.0 |
| | Missing | % | 40.7 | 63.3 | 27.7 | 59.6 | 33.0 |

Indicator variables with less than 5% missing are not displayed.

**Table S3.6**

| Antihypertensive medication | | | Total | No | Yes |
|---|---|---|---|---|---|
| BMI | Present | N | 950 | 160 | 790 |
| | | % | 68.3 | 41.6 | 78.6 |
| | Missing | % | 31.7 | 58.4 | 21.4 |
| SBP | Present | N | 1060 | 182 | 878 |
| | | % | 76.3 | 47.3 | 87.4 |
| | Missing | % | 23.7 | 52.7 | 12.6 |
| DBP | Present | N | 1058 | 182 | 876 |
| | | % | 76.1 | 47.3 | 87.2 |
| | Missing | % | 23.9 | 52.7 | 12.8 |
| HR | Present | N | 715 | 119 | 596 |
| | | % | 51.4 | 30.9 | 59.3 |
| | Missing | % | 48.6 | 69.1 | 40.7 |
| HbA1c | Present | N | 1074 | 193 | 881 |
| | | % | 77.3 | 50.1 | 87.7 |
| | Missing | % | 22.7 | 49.9 | 12.3 |
| FG | Present | N | 1096 | 208 | 888 |
| | | % | 78.8 | 54.0 | 88.4 |
| | Missing | % | 21.2 | 46.0 | 11.6 |
| TC | Present | N | 1067 | 198 | 869 |
| | | % | 76.8 | 51.4 | 86.5 |
| | Missing | % | 23.2 | 48.6 | 13.5 |
| HDL | Present | N | 1056 | 196 | 860 |
| | | % | 76.0 | 50.9 | 85.6 |
| | Missing | % | 24.0 | 49.1 | 14.4 |
| LDL | Present | N | 1053 | 196 | 857 |

| | | | | | |
|---|---|---|---|---|---|
| | | % | 75.8 | 50.9 | 85.3 |
| | Missing | % | 24.2 | 49.1 | 14.7 |
| TC/HDL | Present | N | 1051 | 194 | 857 |
| | | % | 75.6 | 50.4 | 85.3 |
| | Missing | % | 24.4 | 49.6 | 14.7 |
| TG | Present | N | 1052 | 195 | 857 |
| | | % | 75.7 | 50.6 | 85.3 |
| | Missing | % | 24.3 | 49.4 | 14.7 |
| Creat B | Present | N | 1113 | 204 | 909 |
| | | % | 80.1 | 53.0 | 90.4 |
| | Missing | % | 19.9 | 47.0 | 9.6 |
| Creat U | Present | N | 797 | 131 | 666 |
| | | % | 57.3 | 34.0 | 66.3 |
| | Missing | % | 42.7 | 66.0 | 33.7 |
| Alb U | Present | N | 849 | 149 | 700 |
| | | % | 61.1 | 38.7 | 69.7 |
| | Missing | % | 38.9 | 61.3 | 30.3 |
| eGFR | Present | N | 985 | 178 | 807 |
| | | % | 70.9 | 46.2 | 80.3 |
| | Missing | % | 29.1 | 53.8 | 19.7 |
| UACR | Present | N | 824 | 136 | 688 |
| | | % | 59.3 | 35.3 | 68.5 |
| | Missing | % | 40.7 | 64.7 | 31.5 |
| Smoking Behaviour | Present | N | 836 | 144 | 692 |
| | | % | 60.1 | 37.4 | 68.9 |
| | Missing | % | 39.9 | 62.6 | 31.1 |
| eGFR categories | Present | N | 985 | 178 | 807 |
| | | % | 70.9 | 46.2 | 80.3 |
| | Missing | % | 29.1 | 53.8 | 19.7 |
| Albuminuria | Present | N | 824 | 136 | 688 |
| | | % | 59.3 | 35.3 | 68.5 |
| | Missing | % | 40.7 | 64.7 | 31.5 |

Indicator variables with less than 5% missing are not displayed.

**Table S3.7**

| Lipid-lowering medication | | | Total | No | Yes |
|---|---|---|---|---|---|
| BMI | Present | N | 950 | 213 | 737 |
| | | % | 68.3 | 45.5 | 79.9 |
| | Missing | % | 31.7 | 54.5 | 20.1 |
| SBP | Present | N | 1060 | 252 | 808 |

| | | | | | |
|---|---|---|---|---|---|
| | | % | 76.3 | 53.8 | 87.6 |
| | Missing | % | 23.7 | 46.2 | 12.4 |
| DBP | Present | N | 1058 | 250 | 808 |
| | | % | 76.1 | 53.4 | 87.6 |
| | Missing | % | 23.9 | 46.6 | 12.4 |
| HR | Present | N | 715 | 159 | 556 |
| | | % | 51.4 | 34.0 | 60.3 |
| | Missing | % | 48.6 | 66.0 | 39.7 |
| HbA1c | Present | N | 1074 | 247 | 827 |
| | | % | 77.3 | 52.8 | 89.7 |
| | Missing | % | 22.7 | 47.2 | 10.3 |
| FG | Present | N | 1096 | 271 | 825 |
| | | % | 78.8 | 57.9 | 89.5 |
| | Missing | % | 21.2 | 42.1 | 10.5 |
| TC | Present | N | 1067 | 252 | 815 |
| | | % | 76.8 | 53.8 | 88.4 |
| | Missing | % | 23.2 | 46.2 | 11.6 |
| HDL | Present | N | 1056 | 249 | 807 |
| | | % | 76.0 | 53.2 | 87.5 |
| | Missing | % | 24.0 | 46.8 | 12.5 |
| LDL | Present | N | 1053 | 246 | 807 |
| | | % | 75.8 | 52.6 | 87.5 |
| | Missing | % | 24.2 | 47.4 | 12.5 |
| TC/HDL | Present | N | 1051 | 245 | 806 |
| | | % | 75.6 | 52.4 | 87.4 |
| | Missing | % | 24.4 | 47.6 | 12.6 |
| TG | Present | N | 1052 | 246 | 806 |
| | | % | 75.7 | 52.6 | 87.4 |
| | Missing | % | 24.3 | 47.4 | 12.6 |
| Creat B | Present | N | 1113 | 275 | 838 |
| | | % | 80.1 | 58.8 | 90.9 |
| | Missing | % | 19.9 | 41.2 | 9.1 |
| Creat U | Present | N | 797 | 158 | 639 |
| | | % | 57.3 | 33.8 | 69.3 |
| | Missing | % | 42.7 | 66.2 | 30.7 |
| Alb U | Present | N | 849 | 177 | 672 |
| | | % | 61.1 | 37.8 | 72.9 |
| | Missing | % | 38.9 | 62.2 | 27.1 |
| eGFR | Present | N | 985 | 229 | 756 |
| | | % | 70.9 | 48.9 | 82.0 |

| | | | | | |
|---|---|---|---|---|---|
| | Missing | % | 29.1 | 51.1 | 18.0 |
| UACR | Present | N | 824 | 168 | 656 |
| | | % | 59.3 | 35.9 | 71.1 |
| | Missing | % | 40.7 | 64.1 | 28.9 |
| Smoking Behaviour | Present | N | 836 | 187 | 649 |
| | | % | 60.1 | 40.0 | 70.4 |
| | Missing | % | 39.9 | 60.0 | 29.6 |
| eGFR categories | Present | N | 985 | 229 | 756 |
| | | % | 70.9 | 48.9 | 82.0 |
| | Missing | % | 29.1 | 51.1 | 18.0 |
| Albuminuria | Present | N | 824 | 168 | 656 |
| | | % | 59.3 | 35.9 | 71.1 |
| | Missing | % | 40.7 | 64.1 | 28.9 |

Indicator variables with less than 5% missing are not displayed.

**Table S3.8**

| QTc-prolonging medication | | | Total | No | Yes |
|---|---|---|---|---|---|
| BMI | Present | N | 950 | 753 | 197 |
| | | % | 68.3 | 66.8 | 74.9 |
| | Missing | % | 31.7 | 33.2 | 25.1 |
| SBP | Present | N | 1060 | 836 | 224 |
| | | % | 76.3 | 74.2 | 85.2 |
| | Missing | % | 23.7 | 25.8 | 14.8 |
| DBP | Present | N | 1058 | 835 | 223 |
| | | % | 76.1 | 74.1 | 84.8 |
| | Missing | % | 23.9 | 25.9 | 15.2 |
| HR | Present | N | 715 | 558 | 157 |
| | | % | 51.4 | 49.5 | 59.7 |
| | Missing | % | 48.6 | 50.5 | 40.3 |
| HbA1c | Present | N | 1074 | 854 | 220 |
| | | % | 77.3 | 75.8 | 83.7 |
| | Missing | % | 22.7 | 24.2 | 16.3 |
| FG | Present | N | 1096 | 868 | 228 |
| | | % | 78.8 | 77.0 | 86.7 |
| | Missing | % | 21.2 | 23.0 | 13.3 |
| TC | Present | N | 1067 | 842 | 225 |
| | | % | 76.8 | 74.7 | 85.6 |
| | Missing | % | 23.2 | 25.3 | 14.4 |
| HDL | Present | N | 1056 | 833 | 223 |
| | | % | 76.0 | 73.9 | 84.8 |

| | | | | | |
|---|---|---|---|---|---|
| | Missing | % | 24.0 | 26.1 | 15.2 |
| LDL | Present | N | 1053 | 832 | 221 |
| | | % | 75.8 | 73.8 | 84.0 |
| | Missing | % | 24.2 | 26.2 | 16.0 |
| TC/HDL | Present | N | 1051 | 830 | 221 |
| | | % | 75.6 | 73.6 | 84.0 |
| | Missing | % | 24.4 | 26.4 | 16.0 |
| TG | Present | N | 1052 | 832 | 220 |
| | | % | 75.7 | 73.8 | 83.7 |
| | Missing | % | 24.3 | 26.2 | 16.3 |
| Creat B | Present | N | 1113 | 872 | 241 |
| | | % | 80.1 | 77.4 | 91.6 |
| | Missing | % | 19.9 | 22.6 | 8.4 |
| Creat U | Present | N | 797 | 629 | 168 |
| | | % | 57.3 | 55.8 | 63.9 |
| | Missing | % | 42.7 | 44.2 | 36.1 |
| Alb U | Present | N | 849 | 671 | 178 |
| | | % | 61.1 | 59.5 | 67.7 |
| | Missing | % | 38.9 | 40.5 | 32.3 |
| eGFR | Present | N | 985 | 766 | 219 |
| | | % | 70.9 | 68.0 | 83.3 |
| | Missing | % | 29.1 | 32.0 | 16.7 |
| UACR | Present | N | 824 | 654 | 170 |
| | | % | 59.3 | 58.0 | 64.6 |
| | Missing | % | 40.7 | 42.0 | 35.4 |
| Smoking Behaviour | Present | N | 836 | 658 | 178 |
| | | % | 60.1 | 58.4 | 67.7 |
| | Missing | % | 39.9 | 41.6 | 32.3 |
| eGFR categories | Present | N | 985 | 766 | 219 |
| | | % | 70.9 | 68.0 | 83.3 |
| | Missing | % | 29.1 | 32.0 | 16.7 |
| Albuminuria | Present | N | 824 | 654 | 170 |
| | | % | 59.3 | 58.0 | 64.6 |
| | Missing | % | 40.7 | 42.0 | 35.4 |

Indicator variables with less than 5% missing are not displayed.

**Table S3.9**

| **Hypertension** | | | **Total** | **No** | **Yes** |
|---|---|---|---|---|---|
| BMI | Present | N | 950 | 90 | 860 |
| | | % | 68.3 | 42.9 | 72.9 |

| | | | | | |
|---|---|---|---|---|---|
| | Missing | % | 31.7 | 57.1 | 27.1 |
| SBP | Present | N | 1060 | 102 | 958 |
| | | % | 76.3 | 48.6 | 81.2 |
| | Missing | % | 23.7 | 51.4 | 18.8 |
| DBP | Present | N | 1058 | 102 | 956 |
| | | % | 76.1 | 48.6 | 81.0 |
| | Missing | % | 23.9 | 51.4 | 19.0 |
| HR | Present | N | 715 | 61 | 654 |
| | | % | 51.4 | 29.0 | 55.4 |
| | Missing | % | 48.6 | 71.0 | 44.6 |
| HbA1c | Present | N | 1074 | 120 | 954 |
| | | % | 77.3 | 57.1 | 80.8 |
| | Missing | % | 22.7 | 42.9 | 19.2 |
| FG | Present | N | 1096 | 128 | 968 |
| | | % | 78.8 | 61.0 | 82.0 |
| | Missing | % | 21.2 | 39.0 | 18.0 |
| TC | Present | N | 1067 | 121 | 946 |
| | | % | 76.8 | 57.6 | 80.2 |
| | Missing | % | 23.2 | 42.4 | 19.8 |
| HDL | Present | N | 1056 | 120 | 936 |
| | | % | 76.0 | 57.1 | 79.3 |
| | Missing | % | 24.0 | 42.9 | 20.7 |
| LDL | Present | N | 1053 | 120 | 933 |
| | | % | 75.8 | 57.1 | 79.1 |
| | Missing | % | 24.2 | 42.9 | 20.9 |
| TC/HDL | Present | N | 1051 | 119 | 932 |
| | | % | 75.6 | 56.7 | 79.0 |
| | Missing | % | 24.4 | 43.3 | 21.0 |
| TG | Present | N | 1052 | 120 | 932 |
| | | % | 75.7 | 57.1 | 79.0 |
| | Missing | % | 24.3 | 42.9 | 21.0 |
| Creat B | Present | N | 1113 | 126 | 987 |
| | | % | 80.1 | 60.0 | 83.6 |
| | Missing | % | 19.9 | 40.0 | 16.4 |
| Creat U | Present | N | 797 | 71 | 726 |
| | | % | 57.3 | 33.8 | 61.5 |
| | Missing | % | 42.7 | 66.2 | 38.5 |
| Alb U | Present | N | 849 | 82 | 767 |
| | | % | 61.1 | 39.0 | 65.0 |
| | Missing | % | 38.9 | 61.0 | 35.0 |

| | | | | | |
|---|---|---|---|---|---|
| eGFR | Present | N | 985 | 103 | 882 |
| | | % | 70.9 | 49.0 | 74.7 |
| | Missing | % | 29.1 | 51.0 | 25.3 |
| UACR | Present | N | 824 | 74 | 750 |
| | | % | 59.3 | 35.2 | 63.6 |
| | Missing | % | 40.7 | 64.8 | 36.4 |
| Smoking Behaviour | Present | N | 836 | 80 | 756 |
| | | % | 60.1 | 38.1 | 64.1 |
| | Missing | % | 39.9 | 61.9 | 35.9 |
| eGFR categories | Present | N | 985 | 103 | 882 |
| | | % | 70.9 | 49.0 | 74.7 |
| | Missing | % | 29.1 | 51.0 | 25.3 |
| Albuminuria | Present | N | 824 | 74 | 750 |
| | | % | 59.3 | 35.2 | 63.6 |
| | Missing | % | 40.7 | 64.8 | 36.4 |

Indicator variables with less than 5% missing are not displayed.

**Table S3.10**

| Dyslipidemia | | | Total | No | Yes |
|---|---|---|---|---|---|
| BMI | Present | N | 950 | 69 | 881 |
| | | % | 68.3 | 27.4 | 77.4 |
| | Missing | % | 31.7 | 72.6 | 22.6 |
| SBP | Present | N | 1060 | 97 | 963 |
| | | % | 76.3 | 38.5 | 84.6 |
| | Missing | % | 23.7 | 61.5 | 15.4 |
| DBP | Present | N | 1058 | 97 | 961 |
| | | % | 76.1 | 38.5 | 84.4 |
| | Missing | % | 23.9 | 61.5 | 15.6 |
| HR | Present | N | 715 | 60 | 655 |
| | | % | 51.4 | 23.8 | 57.6 |
| | Missing | % | 48.6 | 76.2 | 42.4 |
| HbA1c | Present | N | 1074 | 86 | 988 |
| | | % | 77.3 | 34.1 | 86.8 |
| | Missing | % | 22.7 | 65.9 | 13.2 |
| FG | Present | N | 1096 | 90 | 1006 |
| | | % | 78.8 | 35.7 | 88.4 |
| | Missing | % | 21.2 | 64.3 | 11.6 |
| TC | Present | N | 1067 | 70 | 997 |
| | | % | 76.8 | 27.8 | 87.6 |
| | Missing | % | 23.2 | 72.2 | 12.4 |

| | | | | | |
|---|---|---|---|---|---|
| HDL | Present | N | 1056 | 67 | 989 |
| | | % | 76.0 | 26.6 | 86.9 |
| | Missing | % | 24.0 | 73.4 | 13.1 |
| LDL | Present | N | 1053 | 64 | 989 |
| | | % | 75.8 | 25.4 | 86.9 |
| | Missing | % | 24.2 | 74.6 | 13.1 |
| TC/HDL | Present | N | 1051 | 67 | 984 |
| | | % | 75.6 | 26.6 | 86.5 |
| | Missing | % | 24.4 | 73.4 | 13.5 |
| TG | Present | N | 1052 | 65 | 987 |
| | | % | 75.7 | 25.8 | 86.7 |
| | Missing | % | 24.3 | 74.2 | 13.3 |
| Creat B | Present | N | 1113 | 95 | 1018 |
| | | % | 80.1 | 37.7 | 89.5 |
| | Missing | % | 19.9 | 62.3 | 10.5 |
| Creat U | Present | N | 797 | 47 | 750 |
| | | % | 57.3 | 18.7 | 65.9 |
| | Missing | % | 42.7 | 81.3 | 34.1 |
| Alb U | Present | N | 849 | 53 | 796 |
| | | % | 61.1 | 21.0 | 69.9 |
| | Missing | % | 38.9 | 79.0 | 30.1 |
| eGFR | Present | N | 985 | 79 | 906 |
| | | % | 70.9 | 31.3 | 79.6 |
| | Missing | % | 29.1 | 68.7 | 20.4 |
| UACR | Present | N | 824 | 51 | 773 |
| | | % | 59.3 | 20.2 | 67.9 |
| | Missing | % | 40.7 | 79.8 | 32.1 |
| Smoking Behaviour | Present | N | 836 | 56 | 780 |
| | | % | 60.1 | 22.2 | 68.5 |
| | Missing | % | 39.9 | 77.8 | 31.5 |
| eGFR categories | Present | N | 985 | 79 | 906 |
| | | % | 70.9 | 31.3 | 79.6 |
| | Missing | % | 29.1 | 68.7 | 20.4 |
| Albuminuria | Present | N | 824 | 51 | 773 |
| | | % | 59.3 | 20.2 | 67.9 |
| | Missing | % | 40.7 | 79.8 | 32.1 |

Indicator variables with less than 5% missing are not displayed.

**Table S3.11**

| CVD | Total | No | Yes |
|---|---|---|---|

| | | | | | |
|---|---|---|---|---|---|
| BMI | Present | N | 950 | 590 | 360 |
| | | % | 68.3 | 69.6 | 66.4 |
| | Missing | % | 31.7 | 30.4 | 33.6 |
| SBP | Present | N | 1060 | 663 | 397 |
| | | % | 76.3 | 78.2 | 73.2 |
| | Missing | % | 23.7 | 21.8 | 26.8 |
| DBP | Present | N | 1058 | 662 | 396 |
| | | % | 76.1 | 78.1 | 73.1 |
| | Missing | % | 23.9 | 21.9 | 26.9 |
| HR | Present | N | 715 | 438 | 277 |
| | | % | 51.4 | 51.7 | 51.1 |
| | Missing | % | 48.6 | 48.3 | 48.9 |
| HbA1c | Present | N | 1074 | 667 | 407 |
| | | % | 77.3 | 78.7 | 75.1 |
| | Missing | % | 22.7 | 21.3 | 24.9 |
| FG | Present | N | 1096 | 687 | 409 |
| | | % | 78.8 | 81.0 | 75.5 |
| | Missing | % | 21.2 | 19.0 | 24.5 |
| TC | Present | N | 1067 | 678 | 389 |
| | | % | 76.8 | 80.0 | 71.8 |
| | Missing | % | 23.2 | 20.0 | 28.2 |
| HDL | Present | N | 1056 | 672 | 384 |
| | | % | 76.0 | 79.2 | 70.8 |
| | Missing | % | 24.0 | 20.8 | 29.2 |
| LDL | Present | N | 1053 | 669 | 384 |
| | | % | 75.8 | 78.9 | 70.8 |
| | Missing | % | 24.2 | 21.1 | 29.2 |
| TC/HDL | Present | N | 1051 | 670 | 381 |
| | | % | 75.6 | 79.0 | 70.3 |
| | Missing | % | 24.4 | 21.0 | 29.7 |
| TG | Present | N | 1052 | 668 | 384 |
| | | % | 75.7 | 78.8 | 70.8 |
| | Missing | % | 24.3 | 21.2 | 29.2 |
| Creat B | Present | N | 1113 | 691 | 422 |
| | | % | 80.1 | 81.5 | 77.9 |
| | Missing | % | 19.9 | 18.5 | 22.1 |
| Creat U | Present | N | 797 | 494 | 303 |
| | | % | 57.3 | 58.3 | 55.9 |
| | Missing | % | 42.7 | 41.7 | 44.1 |
| Alb U | Present | N | 849 | 540 | 309 |

| | | | | | |
|---|---|---|---|---|---|
| | | % | 61.1 | 63.7 | 57.0 |
| | Missing | % | 38.9 | 36.3 | 43.0 |
| eGFR | Present | N | 985 | 601 | 384 |
| | | % | 70.9 | 70.9 | 70.8 |
| | Missing | % | 29.1 | 29.1 | 29.2 |
| UACR | Present | N | 824 | 511 | 313 |
| | | % | 59.3 | 60.3 | 57.7 |
| | Missing | % | 40.7 | 39.7 | 42.3 |
| Smoking Behaviour | Present | N | 836 | 510 | 326 |
| | | % | 60.1 | 60.1 | 60.1 |
| | Missing | % | 39.9 | 39.9 | 39.9 |
| eGFR categories | Present | N | 985 | 601 | 384 |
| | | % | 70.9 | 70.9 | 70.8 |
| | Missing | % | 29.1 | 29.1 | 29.2 |
| Albuminuria | Present | N | 824 | 511 | 313 |
| | | % | 59.3 | 60.3 | 57.7 |
| | Missing | % | 40.7 | 39.7 | 42.3 |

Indicator variables with less than 5% missing are not displayed.

**Table S3.12**

| Microvascular complications | | | Total | No | Yes |
|---|---|---|---|---|---|
| BMI | Present | N | 950 | 800 | 150 |
| | | % | 68.3 | 69.4 | 63.3 |
| | Missing | % | 31.7 | 30.6 | 36.7 |
| SBP | Present | N | 1060 | 892 | 168 |
| | | % | 76.3 | 77.4 | 70.9 |
| | Missing | % | 23.7 | 22.6 | 29.1 |
| DBP | Present | N | 1058 | 890 | 168 |
| | | % | 76.1 | 77.2 | 70.9 |
| | Missing | % | 23.9 | 22.8 | 29.1 |
| HR | Present | N | 715 | 589 | 126 |
| | | % | 51.4 | 51.1 | 53.2 |
| | Missing | % | 48.6 | 48.9 | 46.8 |
| HbA1c | Present | N | 1074 | 908 | 166 |
| | | % | 77.3 | 78.8 | 70.0 |
| | Missing | % | 22.7 | 21.2 | 30.0 |
| FG | Present | N | 1096 | 933 | 163 |
| | | % | 78.8 | 80.9 | 68.8 |
| | Missing | % | 21.2 | 19.1 | 31.2 |
| TC | Present | N | 1067 | 906 | 161 |

| | | | | | |
|---|---|---|---|---|---|
| | | % | 76.8 | 78.6 | 67.9 |
| | Missing | % | 23.2 | 21.4 | 32.1 |
| HDL | Present | N | 1056 | 896 | 160 |
| | | % | 76.0 | 77.7 | 67.5 |
| | Missing | % | 24.0 | 22.3 | 32.5 |
| LDL | Present | N | 1053 | 895 | 158 |
| | | % | 75.8 | 77.6 | 66.7 |
| | Missing | % | 24.2 | 22.4 | 33.3 |
| TC/HDL | Present | N | 1051 | 894 | 157 |
| | | % | 75.6 | 77.5 | 66.2 |
| | Missing | % | 24.4 | 22.5 | 33.8 |
| TG | Present | N | 1052 | 894 | 158 |
| | | % | 75.7 | 77.5 | 66.7 |
| | Missing | % | 24.3 | 22.5 | 33.3 |
| Creat B | Present | N | 1113 | 933 | 180 |
| | | % | 80.1 | 80.9 | 75.9 |
| | Missing | % | 19.9 | 19.1 | 24.1 |
| Creat U | Present | N | 797 | 667 | 130 |
| | | % | 57.3 | 57.8 | 54.9 |
| | Missing | % | 42.7 | 42.2 | 45.1 |
| Alb U | Present | N | 849 | 717 | 132 |
| | | % | 61.1 | 62.2 | 55.7 |
| | Missing | % | 38.9 | 37.8 | 44.3 |
| eGFR | Present | N | 985 | 817 | 168 |
| | | % | 70.9 | 70.9 | 70.9 |
| | Missing | % | 29.1 | 29.1 | 29.1 |
| UACR | Present | N | 824 | 690 | 134 |
| | | % | 59.3 | 59.8 | 56.5 |
| | Missing | % | 40.7 | 40.2 | 43.5 |
| Smoking Behaviour | Present | N | 836 | 696 | 140 |
| | | % | 60.1 | 60.4 | 59.1 |
| | Missing | % | 39.9 | 39.6 | 40.9 |
| eGFR categories | Present | N | 985 | 817 | 168 |
| | | % | 70.9 | 70.9 | 70.9 |
| | Missing | % | 29.1 | 29.1 | 29.1 |
| Albuminuria | Present | N | 824 | 690 | 134 |
| | | % | 59.3 | 59.8 | 56.5 |
| | Missing | % | 40.7 | 40.2 | 43.5 |

Indicator variables with less than 5% missing are not displayed.

Table S4. Sensitivity analysis comparing associations with SCA from conditional and unconditional (adjusted for sex and age) logistic regression models.

| Clinical characteristic | Univariable OR of SCA | | Multivariable OR of SCA | |
|---|---|---|---|---|
| | Conditional LR | Unconditional LR | Conditional LR | Unconditional LR |
| Sex (matching variable) | n.a. | # | n.a. | 1.05 (0.75-1.48) |
| Age (matching variable) | n.a. | # | n.a. | 0.99 (0.98-1.01) |
| Smoking behavior (%) | | | | |
| Never | *Reference* | *Reference* | *Reference* | *Reference* |
| Former | 0.97 (0.64-1.46) | 0.96 (0.64-1.44) | 0.84 (0.53-1.31) | 0.86 (0.56-1.32) |
| Current | 1.81 (1.17-2.80)** | 1.82 (1.18-2.81)** | 1.62 (0.99-2.71) | 1.70 (1.05-2.74)* |
| BMI (kg/m$^2$) | 1.03 (1.00-1.06) | 1.03 (1.00-1.06) | 1.02 (0.98-1.05) | 1.02 (0.98-1.05) |
| HbA1c (mmol/mol) | 1.00 (1.00-1.00) | 1.00 (1.00-1.00) | 1.00 (0.99-1.00) | 1.00 (1.00-1.00) |
| Fasting glucose (mmol/mol) | 1.08 (1.01-1.14)* | 1.07 (1.01-1.14)* | 1.08 (1.01-1.16)* | 1.07 (1.01-1.14)* |
| TC/HDL ratio | 1.19 (1.06 -1.33)** | 1.18 (1.06 -1.33)** | 1.17 (1.03-1.34)* | 1.18 (1.04-1.35)* |
| eGFR (ml/min) | | | | |
| Normal/high (>90) | *Reference* | *Reference* | *Reference* | *Reference* |
| Mildly decreased (<90 - ≥60) | 1.08 (0.61-1.92) | 1.01 (0.59-1.74) | 1.11 (0.59-2.09) | 1.04 (0.58-1.87) |
| Moderately decreased (<60 - ≥30) | 1.83 (0.99-3.37) | 1.70 (0.94-3.08) | 1.53 (0.77-3.05) | 1.39 (0.73-2.67) |
| Severely decreased (<30) | 3.12 (1.49-6.57)** | 2.62 (1.33-5.14)** | 2.19 (0.94-5.08) | 1.80 (0.83-3.90) |
| Albuminuria (mg/mmol) | | | | |
| Normal/mild (<3) | *Reference* | *Reference* | *Reference* | *Reference* |
| Moderate (≥3 - ≤ 30) | 2.95 (2.02-4.31)** | 2.83 (1.97-4.05)** | 2.77 (1.84-4.16)** | 2.64 (1.79-3.89)** |
| Severe (>30) | 4.49 (2.39-8.44)** | 4.23 (2.35-7.61)** | 2.96 (1.44-6.08)** | 2.74 (1.41-5.30)** |
| Glucose-lowering medication | | | | |
| No medication | *Reference* | *Reference* | *Reference* | *Reference* |
| Oral only | 1.14 (0.80-1.62) | 1.01 (0.73-1.40) | 1.06 (0.69-1.64) | 1.12 (0.76-1.67) |
| Insulin only | 3.21 (1.63-6.23)** | 2.94 (1.58-5.48)** | 1.97 (0.88-4.39) | 2.34 (1.14-4.77)* |
| Oral & insulin | 1.59 (0.99-2.53) | 1.41 (0.92-2.15) | 0.86 (0.48-1.54) | 0.96 (0.56-1.65) |
| QTc prolonging medication (yes) | 1.14 (0.80-1.62) | 1.11 (0.78-1.57) | 1.02 (0.69-1.52) | 1.07 (0.73-1.57) |
| Hypertension (yes) | 1.59 (1.02-2.48)* | 1.46 (0.95-2.24) | 1.33 (0.79-2.25) | 1.36 (0.83-2.22) |
| Dyslipidemia (yes) | 0.65 (0.44-0.96)* | 0.67 (0.48-0.93)* | 0.53 (0.33-0.86)* | 0.51 (0.33-0.80)* |
| History of CVD (yes) | 2.05 (1.53-2.73)** | 1.94 (1.46-2.57)** | 1.72 (1.23-2.40)** | 1.55 (1.13-2.13)** |

| | | | | |
|---|---|---|---|---|
| Microvascular complications (yes) | 1.86 (1.34-2.67)** | 1.79 (1.28-2.49)** | 1.44 (0.96-2.17) | 1.36 (0.93-1.99) |

### the estimates for age and sex differ slightly in the various unconditional univariable models (adjusted for matching variables age and sex), but are all around 1 and insignificant.

Data are presented as pooled odds ratios (OR (95% CI)) from conditional and unconditional logistic regression models after multiple imputation. The univariable unconditional model was adjusted for matching variables sex and age. In the multivariable model all variables were entered simultaneously (including sex and age for the unconditional model).

LR, logistic regresion; N.a., not applicable; BMI, body mass index; HbA1c, hemoglobin A1c; TC, total cholesterol; HDL, high density lipo-protein; eGFR, estimated glomerular filtration rate; QTc, heart rate-corrected QT interval;  CVD, Cardiovascular disease.

* Significant at the $p < 0.05$ level.

** Significant at the $p < 0.01$ level.

Table S5. Sensitivity analyses assessing the influence of multiple imputation by comparing the univariable associations with SCA from original with imputed datasets.

| **Clinical characteristics with missing values** | **Univariable OR of SCA** | |
|---|---|---|
| | **Original dataset** | **Multiple imputation datasets** |
| Smoking behavior (%) | | |
| Never | *Reference* | *Reference* |
| Former | 0.88 (0.53-1.45) | 0.97 (0.64-1.46) |
| Current | 1.54 (0.91-2.60) | 1.81 (1.17-2.80)** |
| BMI (kg/m$^2$) | 1.01 (0.98-1.05) | 1.03 (1.00-1.06) |
| HbA1c (mmol/mol) | 1.00 (1.00-1.00) | 1.00 (1.00-1.00) |
| Fasting glucose (mmol/mol) | 1.08 (1.01-1.15)** | 1.08 (1.01-1.14)* |
| TC/HDL ratio | 1.17 (1.04-1.32)** | 1.19 (1.06 -1.33)** |
| eGFR (ml/min) | | |
| Normal/high (>90) | *Reference* | *Reference* |
| Mildly decreased (<90 - ≥60) | 1.49 (0.78-2.86) | 1.08 (0.61-1.92) |
| Moderately decreased (<60 - ≥30) | 2.57 (1.26-5.24)** | 1.83 (0.99-3.37) |
| Severely decreased (<30) | 4.60 (1.83-11.61)** | 3.12 (1.49-6.57)** |
| Albuminuria (mg/mmol) | | |
| Normal/mild (<3) | *Reference* | *Reference* |
| Moderate (≥3 - ≤ 30) | 2.53 (1.60-4.01)** | 2.95 (2.02-4.31)** |
| Severe (>30) | 4.33 (1.94-9.68)** | 4.49 (2.39-8.44)** |

Data are presented as odds ratios (OR (95% CI)) from univariable conditional logistic regression models from original (pairwise exclusion of missing values) and imputed datasets.
BMI, body mass index; HbA1c, hemoglobin A1c; TC, total cholesterol; HDL, high density lipo-protein; eGFR, estimated glomerular filtration rate; QTc, heart rate-corrected QT interval; CVD, Cardiovascular disease.
* Significant at the p < 0.05 level.
** Significant at the p < 0.01 level.
